# *Schistosoma mansoni* infection alters the host pre-vaccination environment resulting in blunted Hepatitis B vaccination immune responses

**DOI:** 10.1101/2023.02.24.23284435

**Authors:** Roshell Muir, Talibah Metcalf, Slim Fourati, Yannic Bartsch, Jacqueline Kyosiimire Lugemwa, Glenda Canderan, Galit Alter, Enoch Muyanja, Brenda Okech, Teddy Namatovu, Irene Namara, Annemarie Namuniina, Ali Ssetaala, Juliet Mpendo, Annet Nanvubya, Paul Kato Kitandwe, Bernard S. Bagaya, Noah Kiwanuka, Jacent Nassuna, Victoria Menya Biribawa, Alison M. Elliott, Claudia J. de Dood, William Senyonga, Priscilla Balungi, Pontiano Kaleebu, Yunia Mayanja, Mathew Odongo, Pat Fast, Matt A. Price, Paul L.A.M. Corstjens, Govert J. van Dam, Anatoli Kamali, Rafick Pierre Sekaly, Elias K Haddad

**Affiliations:** Division of Infectious Diseases and HIV Medicine, Department of Medicine, Drexel University College of Medicine, Philadelphia, Pennsylvania, USA; PATRU, School of Medicine, Emory University, Atlanta, GA, USA; Ragon Institute of MGH, MIT, and Harvard, Cambridge, MA, USA; MRC/UVRI and LSHTM Uganda Research Unit, Entebbe, Uganda; Department of Medicine, Allergy and Immunology, University of Virginia, Charlottesville, VA, USA; UVRI-IAVI HIV Vaccine Program, Entebbe, Uganda; Department of Immunology and Molecular Biology, School of Biomedical Sciences, Makerere University, College of Health Sciences, Kampala-Uganda; Department of Epidemiology and Biostatistics, School of Public Health, Makerere University, College of Health Sciences, Kampala-Uganda; Department of Clinical Research, London School of Hygiene and Tropical Medicine, London, UK; Department of Cell and Chemical Biology, Leiden University Medical Center, Leiden, Netherlands; International AIDS Vaccine Initiative, New York, NY, USA; Pediatric Infectious Diseases, Stanford University School of Medicine, Palo Alto, CA, USA; Department of Epidemiology and Biostatistics, University of California at San Francisco, San Francisco, USA; Department of Cell and Chemical Biology, Leiden University Medical Center, Leiden, the Netherlands; Department of Parasitology, Leiden University Medical Center, Leiden, the Netherlands; IAVI, New York, New York, USA, and Nairobi, Kenya

## Abstract

The impact of endemic infections on protective immunity is critical to inform vaccination strategies. In this study, we assessed the influence of *Schistosoma mansoni* infection on host responses in a Ugandan fishing cohort given a Hepatitis B (HepB) vaccine. Concentrations of schistosome-specific circulating anodic antigen (CAA) pre-vaccination showed a significant bimodal distribution associated with HepB titers, which were lower in individuals with high CAA. We established that participants with high CAA had significantly lower frequencies of circulating T follicular helper (cTfh) subpopulations pre- and post-vaccination and higher regulatory T cells (Tregs) post-vaccination. Polarization towards higher frequencies of Tregs: cTfh cells can be mediated by changes in the cytokine environment favoring Treg differentiation. In fact, we observed higher levels of CCL17 and soluble IL-2R pre-vaccination (important for Treg recruitment and development), in individuals with high CAA that negatively associated with HepB titers. Additionally, alterations in pre-vaccination monocyte function correlated with HepB titers, and changes in innate-related cytokines/chemokine production were associated with increasing CAA concentration. We report, that by influencing the immune landscape, schistosomiasis has the potential to modulate immune responses to HepB vaccination. These findings highlight multiple *Schistosoma*-related immune associations that could explain abrogated vaccine responses in communities with endemic infections.

**Author Summary:** Schistosomiasis drives host immune responses for optimal pathogen survival, potentially altering host responses to vaccine-related antigen. Chronic schistosomiasis and co-infection with hepatotropic viruses are common in countries where schistosomiasis is endemic. We explored the impact of *Schistosoma mansoni* (*S. mansoni*) infection on Hepatitis B (HepB) vaccination of individuals from a fishing community in Uganda. We demonstrate that high schistosome-specific antigen (circulating anodic antigen, CAA) concentration pre-vaccination, is associated with lower HepB antibody titers post-vaccination. We show higher pre-vaccination levels of cellular and soluble factors in instances of high CAA that are negatively associated with HepB antibody titers post-vaccination, which coincided with lower frequencies of circulating T follicular helper cell populations (cTfh), proliferating antibody secreting cells (ASCs), and higher frequencies of regulatory T cells (Tregs). We also show that monocyte function is important in HepB vaccine responses, and that high CAA is associated with alterations in the early innate cytokine/chemokine microenvironment. Our findings suggest that in individuals with high CAA and likely high worm burden, schistosomiasis creates and sustains an environment that is polarized against optimal host immune responses to the vaccine, which puts many endemic communities at risk for infection against HepB and other diseases that are preventable by vaccines.

## Introduction

While vaccines have reshaped public health and saved millions of lives from the threat of many infectious diseases (1), inadequate immune responses to vaccination remain a challenge, with a range of contributary causes, such as chronic illnesses, including those stemming from helminth worm infections (2–4). Schistosomiasis is a neglected tropical disease (NTD) caused by parasitic flatworms from three main *Schistosoma* spp. that often develops into a chronic infection in endemic populations, with 90% of all infections found in sub-Saharan Africa. An estimated four million people in Uganda are infected with *S. mansoni* and about 55% of the population is at risk of infection (5, 6). Currently, treatment with Praziquantel (PZQ) is the only safe and effective protocol against infection with *Schistosoma* spp. and control of Schistosomiasis in endemic regions involves periodic administration of PZQ to communities (7).

Chronic schistosomiasis with hepatitis B virus (HBV) and hepatitis C virus (HCV) co-infection is common in countries where schistosomiasis is endemic. There is also evidence that *S. mansoni* co-infection with hepatotropic viruses may exacerbate resultant hepatic pathology (8). In Uganda, more than 1.4 million adults are chronically infected with hepatotropic viruses, and the prevalence of HBV is approximately 8.5% (9, 10) with an increased incidence in the Lake Victoria fishing communities (11). While vaccination against Hepatitis B (HepB) is highly effective in the prevention of infection, there are still ∼ 800,000 deaths attributable to acute and chronic HepB disease annually (12). Furthermore, 5-10% of individuals do not mount an effective antibody response to HepB vaccination (13). There is a need for improved understanding of the immunological responses during vaccination to ensure effective responses in compromised host immune systems.

*Schistosoma* spp. induces strong host T helper 2 (Th2) responses, but also go on to induce regulatory T cell (Treg) responses which in turn curbs T helper immunity (14, 15). Specifically, the chronic phase develops due to continual exposure to the soluble egg antigen (SEA) of schistosome eggs in the host tissues, driving immune-pathology and progressing to granuloma formation, characterized by an increase in lymphocytes (CD4^+^ T cells) producing T-helper-2 (Th2) cytokines such as interleukins (IL) -4, -5, and -13. This heightened Th2 response becomes suppressed over time to induce tolerance, which is the major influence on worm survival in the host (16, 17). Upregulation of these responses could potentially inhibit the T helper responses induced by vaccination, such as T follicular helper (Tfh) responses which are critical for B cell help (18, 19), and foster conditions that cause alterations of vaccine-induced antibody responses. Studies showing the effects of *Schistosoma* infection on the modulation of immune responsiveness to concomitant diseases and immunization are not without controversy; however, the consensus is this infection can diminish or alter immune responses to non-schistosome antigens. These responses may lead to either beneficial or detrimental outcomes for the host. In the case of vaccine responses, animal models of malaria (20), bacteria (21) and HepB (22) show reduced protective effects; and human vaccination studies have shown lower levels of vaccine-specific antibody in *Schistosoma*-infected children (23, 24), and in HepB vaccination of adults (25, 26). However, the impact of *S. mansoni* infection and worm burden on host immune response to vaccines, and to HepB vaccination, is still poorly understood; and the underlying actions of immune mediators affecting these responses need to be further elucidated to identify strategies to optimize vaccination effectiveness.

Detection of CAA (Circulating Anodic Antigen, a worm antigen regurgitated into the bloodstream of the host) in serum indicates an ongoing infection, implying the presence of living *Schistosoma* worms. CAA is produced at a steady level and although worm pairs do produce eggs, worms do not multiply and there is a direct association of CAA serum level with worm burden. It was estimated that an amount of 1-10 pg/mL blood corresponds to a single healthy worm pair (27, 28). In this study, we characterized soluble and cellular immune factors at pre- and post-immunization timepoints with a HepB vaccine in PZQ-treated individuals with high and low CAA and explored mechanisms which may be responsible for a blunted vaccine response in individuals with high CAA and likely a high worm burden. We demonstrated that *S. mansoni* infection is associated with significantly lower HepB vaccine-specific antibody responses and showed these responses coincided with alterations in cytokines and chemokines that are important for Tfh and Treg cell recruitment and function. We propose that a high CAA concentration due to a high *S. mansoni* worm burden pre-vaccination, can reshape the baseline environment, and modulate the response to vaccines.

## Results

### Pre-vaccination *S. mansoni* infection negatively impacts Hepatitis B antibody titers post-vaccination

Seventy-five participants for the present study were recruited in Uganda among fishing communities on the northern shores of Lake Victoria, where schistosomiasis is endemic and various samples were collected at multiple timepoints for subsequent study analyses (Fig 1, S1 Table). HepB antibody titers (anti-HBs) were determined in the serum at D0, M7 (1 month post-2^nd^ boost) and M12 (6 months post-2^nd^ boost) to look at levels pre-vaccine, as well as short-term and longer-term post-vaccination antibody responses after completion of the immunization series (Fig 1 and S1 Table). A circulating anodic antigen (CAA) detection assay (29–32) was also determined pre-vaccination (D0) in the serum of enrolled participants (Fig 1 and S1 Table). The presence of CAA indicates an ongoing *S. mansoni* infection, with CAA concentration closely linked to worm burden (27, 28) and those found to be infected were treated with PZQ at day 12 (D12) post-vaccination (Fig 1). The results show that CAA values displayed a bimodal distribution. The cutoff separating the two modes was estimated by maximum-likelihood which revealed that participants could be separated into those with a CAA concentration of <36 pg/mL (low or no worm burden); and those with a CAA concentration of ≥36 pg/mL (higher worm burden) (Fig 2A). Importantly, CAA values from these two groups of participants were significantly associated with HepB titers measured at month 7 (M7), where low CAA was associated with a high titer, and high CAA with a low titer (log10 fold change (FC)= -0.43, 95% confidence interval (CI) [-0.053, -0.81]; P*=*0.028) (Fig 2B). The same trend between HepB titers and the two CAA groups was observed after adjusting for age and sex (log10FC= -0.38, 95% CI [0.059, -0.81]; P=0.089).

**Fig 1.**
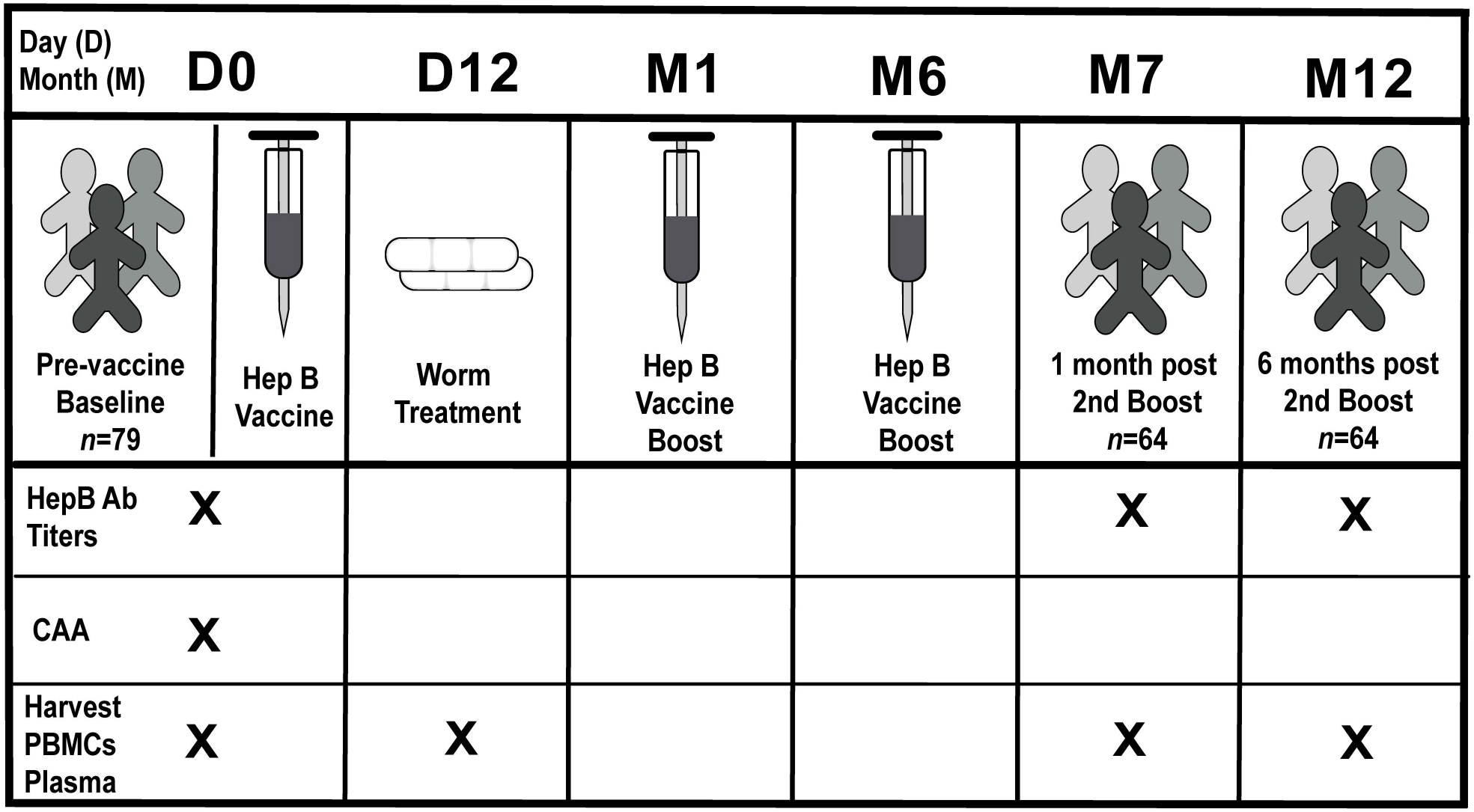
Clinical study design. Participants enrolled in the clinical study (n =79, four donor samples were unavailable for analysis in this study) were vaccinated at baseline [(pre-vaccination) day 0 (D0)] and received two boosters at month 1 (M1) and month 6 (M6). Individuals provided sera samples at D0, month 7 post-vaccination (M7) (one month post-booster 2) and month 12 post-vaccination (M12) (six months post-booster 2), which were used to screen for Hepatitis B (HepB) antibody titers. Enrollees were treated at day 12 post-vaccination (D12) with Praziquantel (PZQ) as appropriate, for egg counts detected in stool samples which were collected at D0 (and day 3 and/or day 7). Sera samples at D0 were also used to evaluate for schistosome-specific antigen (circulating anodic antigen, CAA). PMBCs and plasma were also collected at D0, D12, M7 and M12 for subsequent analysis. X denotes corresponding samples collected at that timepoint.

**Fig 2.**
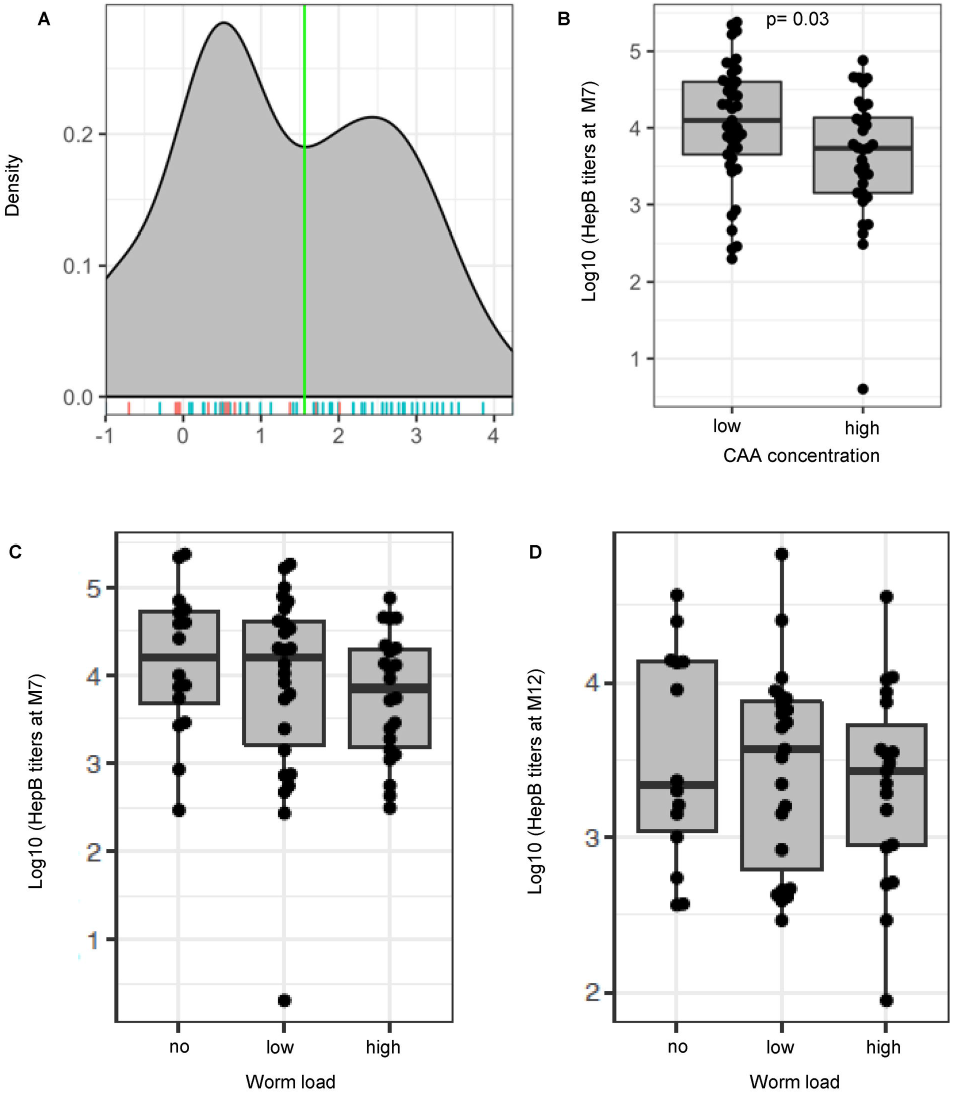
Higher CAA concentration pre-vaccination is associated with a reduction in Hepatitis B vaccine titers. (A) Density of Schistosome-specific antigen values (CAA [pg/ml]) analyzed in serum samples from participants pre-vaccination (D0). A binomial distribution was fitted to the CAA values and a maximum-likelihood was used to identify the optimum cutoff separating the two modes of the CAA values. Two groups of participants were then identified based on CAA values, low CAA (<36pg/mL CAA), n =41; and high CAA (≥36pg/mL CAA), n =33. Male (blue line), n =53; Female (red line), n =21 (B) Hepatitis B (HepB) titers (log pg/mL) determined by commercial immunoassays for individuals at M7 post-vaccination plotted to compare low and high CAA. Participants were further defined based on CAA concentration values using methodology from the CAA assay that allowed detection of samples as low as 3pg/mL: non-infected [no] (<3pg/mL CAA), low CAA [low] (3-100 pg/mL CAA), and high CAA [high] (>100pg/mL CAA) (C) HepB titers for individuals at Month 7 post-vaccination, non-infected, n =16, low CAA, n =26 and high CAA, n =22 and (D) Month 12 post-vaccination, non-infected, n =14, low CAA, n =23, and high CAA, n =19 plotted to compare CAA. Boxplots show median values (horizontal line), interquartile range (box) and 95% confidence interval (whiskers).

In acknowledging the role *S. mansoni* infection and worm burden could play in the modulation of host immunity, whether the individuals at the time of the study were uninfected or infected, and the level of *S. mansoni* worm burden, is important in data interpretation. For this reason, individuals were further stratified into three groups using methodology from the CAA assay that allowed detection of samples as low as 3pg/mL: non-infected (no) (CAA <3 pg/mL), low CAA concentration (low) (CAA 3-100 pg/mL) and high CAA concentration (high) (CAA >100 pg/mL) (S1 Table) (33). Association of these three groups with vaccine responses indicated higher HepB titers in non-infected individuals, followed by those with low CAA and high CAA (Fig 2C and 2D). Overall, these data show an association between CAA concentration pre-vaccination and vaccine-specific responses, specifically that that a higher CAA concentration and likely higher worm burden, can negatively influence post-vaccination antibody titers.

### Plasma cytokines/chemokines involved in lymphocyte migration and activation are significantly higher in *S. mansoni* infection pre-vaccination and persist at month 12 post-vaccination

To elucidate any patterns of cytokine/chemokine production between infected groups pre-vaccination and post-vaccination, plasma samples from non-infected and *S. mansoni*-infected individuals were analyzed using multiplex cytokine/chemokine platform of more than 65 soluble factors. We identified that pre-vaccination systemic levels of CCL17 (no vs. low P=0.002, no vs. high P<0.0001), CCL19 (no vs. low P=0.004, no vs. high P<0.0001, low vs. high P=0.040), CXCL9 (no vs. low P=0.012, no vs. high P<0.0001, low vs. high P=0.002) (Fig 3A-3C, S2 Table), and several other cytokines and chemokines implicated in lymphocyte recruitment and activation (S1A-S1F Fig, S2 Table), were significantly higher in individuals with low and high CAA compared to non-infected at D0. Of interest, the levels of CCL17 (no vs. high P=0.012), CCL19 (no vs. high P=0.040) and CXCL9 (no vs. high P=0.004, low vs. high P=0.031) remained significantly elevated in individuals with high CAA at M12 post-vaccination (Fig 3A-3C, S2 Table). Furthermore, principal component analysis (PCA) analysis shows that cytokine/chemokine levels pre-vaccination, cluster separately from post-vaccination time points, suggesting an association between HepB vaccination and major changes in circulating cytokines/chemokines that are sustained over time (up to 6 months after the last immunization, Fig 2D). The relationship between alterations in these cytokines/chemokines and the levels of HepB titers, was evaluated to identify biomarkers associated with a stronger humoral response to vaccination. Pre-vaccination (t=-3.01, P=0.004) and M12 post-vaccination (t=-2.40, P=0.020) levels of CCL17 showed a negative correlation with HepB titers at M12 (S2A and S2B Fig, S3 Table). While levels of soluble IL-2R (sIL-2R) were not different in *S. mansoni* infection compared to non-infected (S2C Fig and S2 Table), sIL-2R levels pre-vaccination (t=-2.22, P=0.032) and M12 post-vaccination (t=-2.95, P=0.005) were also negatively correlated with HepB titers at M12 (S2D and S2E Fig, S3 Table). These observations suggest a high worm burden in *S. mansoni* infection could be associated with alterations of the host cytokine/chemokine response to vaccination and implicate CCL17 and potentially sIL-2R, known to attract both effector and regulatory T cells (34–38).

**Fig 3.**
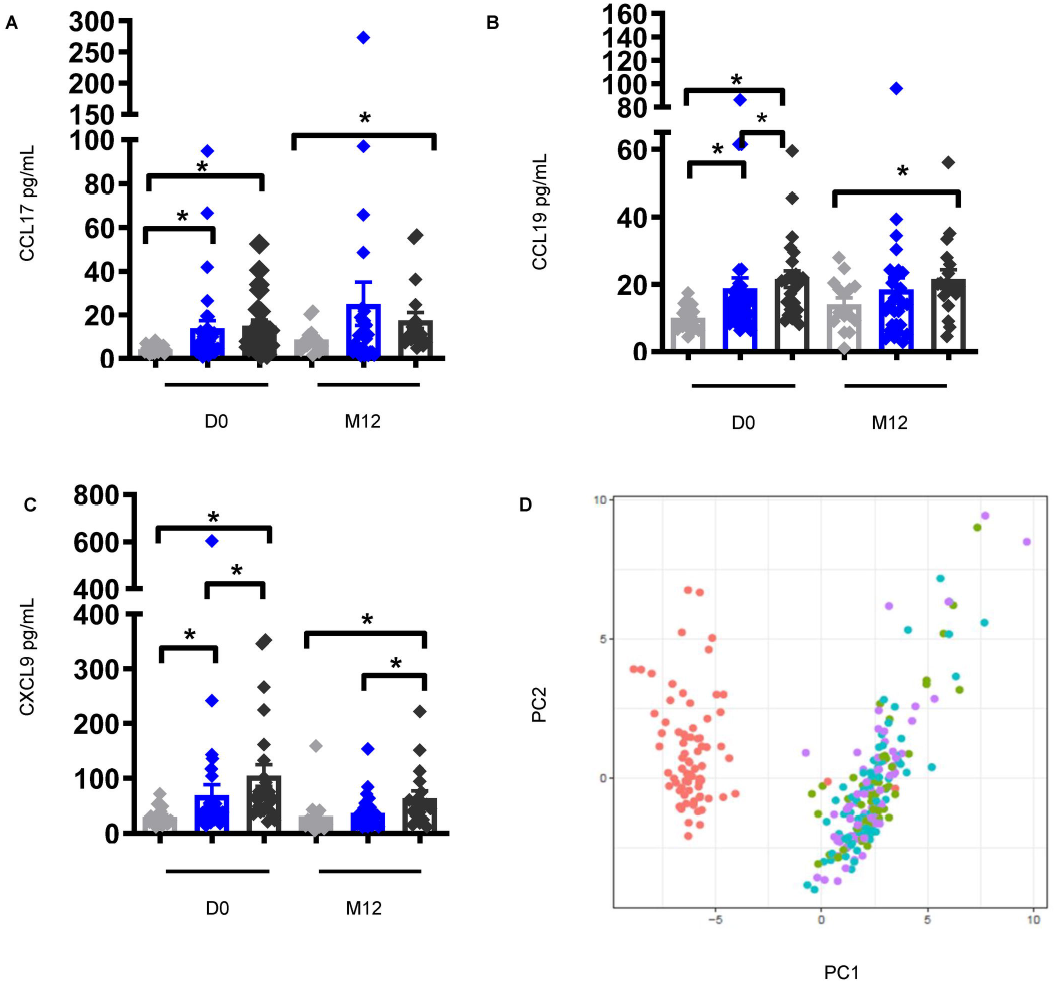
Elevated levels of plasma cytokines/chemokines involved in lymphocyte cell migration and activation pre-vaccination in individuals with *S. mansoni* infection persist at month 12 post-vaccination. (A) CCL17, (B) CCL19, and (C) CXCL9 levels in the plasma of non-infected individuals pre-vaccination (D0), n =19, low CAA, n =32, and high CAA, n =24, and M12 post-vaccination, n =15, low CAA, n =29, and high CAA, n =19. Data shown as ± SEM. * P ≤ 0.05. Wilcoxon rank-sum test performed on non-infected vs. low CAA, or non-infected vs. high CAA, or low CAA vs. high CAA within each time point D0 or M12. Non-infected-light grey, low CAA - blue, and high CAA - dark grey. (D) Principal component analysis of plasma cytokines/chemokines pre-vaccination and after Hepatitis B vaccination was conducted and the first (PC1) and second (PC2) principal components were used to plot samples based on their plasma cytokines/chemokines profiles. Each dot corresponds to a sample and colors denote the timepoint the sample was collected: D0-red, D3-green, D7-blue, and M12-purple.

### Frequencies of circulating T follicular helper cell populations are significantly lower pre-vaccination which is sustained post-vaccination in *S. mansoni* infection and coincides with higher frequencies of regulatory T cells in individuals with high CAA concentration

Cellular immunological changes influenced by the cytokine environment are key in modulating effective vaccine-specific humoral responses. We assessed frequencies of circulating T follicular helper (cTfh) populations (39), critically involved in providing B cell help in infection and vaccination (40–42), from the PBMCs of non-infected and *S. mansoni*-infected individuals by flow cytometry (S3 Fig). Frequencies of cTfh1 (low vs. high P=0.020) and cTfh2 cells (no vs. high P=0.012, low vs. high P=0.014) pre-vaccination (Fig 4A and 4B, S4 Table), were significantly lower in individuals with high CAA; and cTfh1 cells remained significantly lower at M7 (low vs. high P=0.031) and M12 (no vs. high P=0.030, low vs. high P=0.004) post-vaccination (Fig 4A, S4 Table). While differences between non-infected and *S. mansoni*-infected groups for cTfh17 cells pre-vaccination did not reach statistical significance, they were significantly lower in *S. mansoni*-infected individuals at M12 post-vaccination (no vs. low P= 0.024, no vs. high P=0.006) (Fig 4C, S4 Table). Importantly, these alterations were specific to cTfh cells, as no statistically significant changes between groups were observed at any of the time points for non-cTfh cells (Fig 4D). By contrast, frequencies of CXCR5^-^ regulatory T cells (Tregs), that were similar between non-infected and *S. mansoni*-infected groups pre-vaccination, were significantly higher at M7 (no vs. high P=0.039) and M12 (no vs. high P=0.047) post-vaccination in individuals with high CAA (Fig 4E, S4 Table). These results suggest that frequencies of cTfh cells, which are important for instructing B cells to produce antibodies, are significantly altered in instances of high worm burden, and the concomitant increase in Tregs supports the theory that worm burden could have an impact on the interplay between these two subsets.

**Fig 4.**
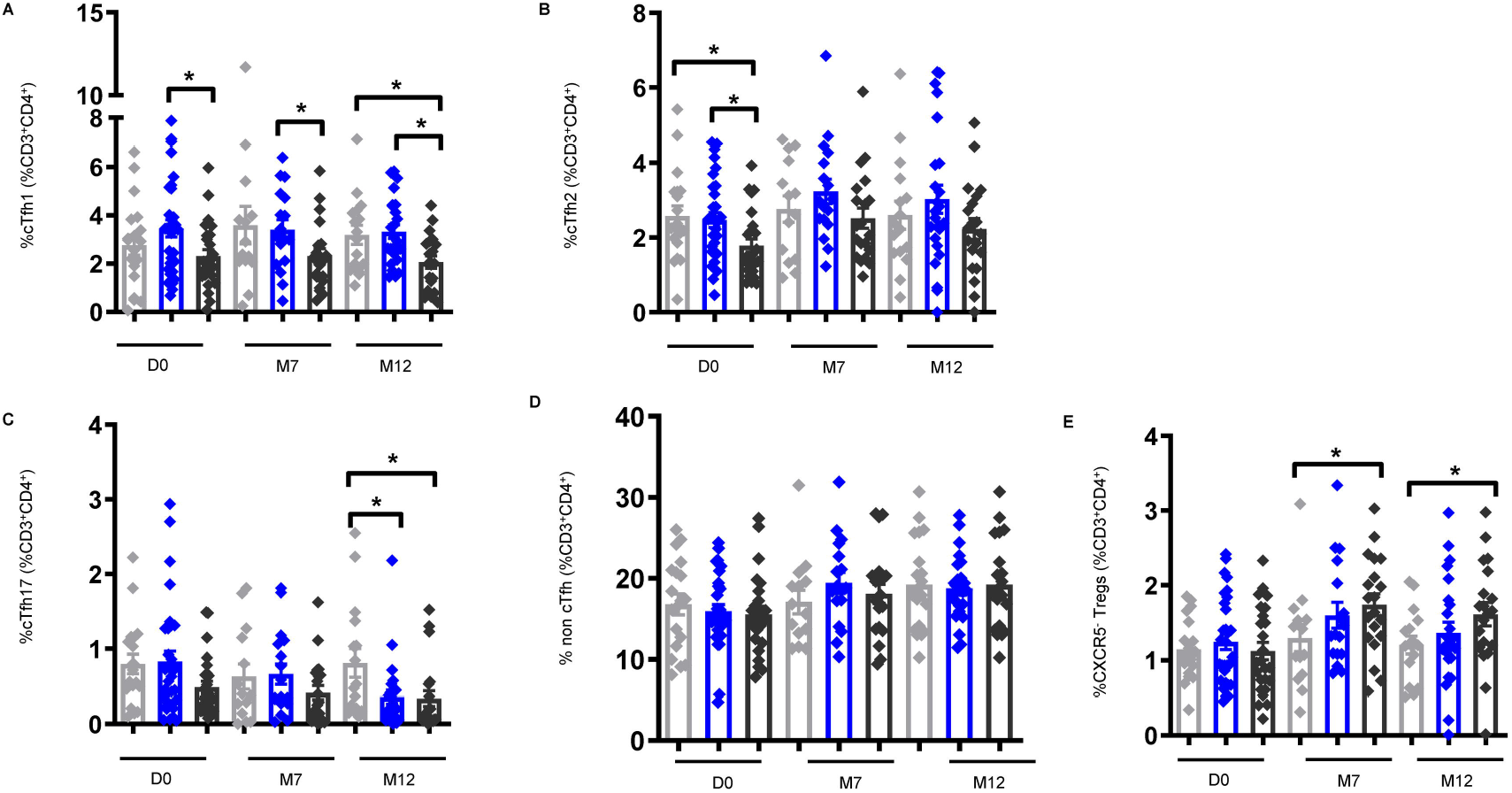
Frequencies of circulating follicular helper (cTfh) cells are lower pre- and post-vaccination in *S. mansoni* infection with concurrent higher frequencies of regulatory T cells (Tregs) in individuals with high CAA concentration. Frequencies of CD3^+^CD4^+^CD45RA^-^CD25^-^CXCR5^+^, populations: (A) cTfh1 [CXCR3^+^], (B) cTfh2 [CXCR3^-^CCR6^-^], (C) cTfh17 [CXCR3^-^CCR6^+^], (D) non cTfh [CD3^+^CD4^+^CD45RA^-^CD25^-^CXCR5^-^] and (E) CXCR5^-^Tregs [CD3^+^CD4^+^CD45RA^-^CD127^-^ CD25^+^Foxp3^+^CXCR5^-^] were identified by flow cytometry of PBMCs from individuals pre-vaccination (D0) non-infected, n =19, low CAA, n =31, and high CAA, n =25, M7 post-vaccination non-infected, n =14, low CAA, n =17, and high CAA, n =20, and M12 post-vaccination non-infected, n =16, low CAA, n =24, and high CAA, n =20. Data shown as ± SEM. *P ≤ 0.05. Wilcoxon rank-sum test performed on non-infected vs. low CAA, or non-infected vs. high CAA, or low CAA vs. high CAA for each time point separately D0, M7, or M12. Non-infected-light grey, low CAA - blue, and high CAA - dark grey.

### Antibody-secreting B cells are significantly lower in instances of high worm burden and is associated with Hepatitis B antibody titers at month 12

Cellular changes were further examined in response to *S. mansoni* infection and HepB vaccination by looking at the frequencies of activated B cells (ABC) and antigen-specific antibody-secreting B cells (ASC) (43) (S4 Fig). Frequencies of ABCs (no vs. high P=0.048) and IgG^+^ABCs (no vs. high P=0.015) were found to be significantly higher pre-vaccination in individuals with high CAA, with similar results at M7 (ABC, no vs. high P=0.039; IgG^+^ABCs, no vs. high P=0.032) and M12 (ABC, no vs. high P=0.040; IgG^+^ABCs, no vs. high P=0.033) post-vaccination (S5A and S5B Fig, S5 Table), indicative of a hyperactivated inflammatory B cell response (41). Of importance and in sharp contrast, lower frequencies of KI67^+^ASC (no vs. high P=0.040), IgG^+^ASC (no vs. high P=0.046), and IgA^+^ASC (no vs. high P=0.029) populations were observed in individuals with high CAA at M12 post-vaccination (Fig 5A-5C). Furthermore, assessment of the correlation between ASCs and HepB titers showed a positive correlation between IgA^+^ASCs and HepB titers at M12 (t=2.10, P=0.041) (S5C Fig and S6 Table). These results imply that an elevated worm burden can influence humoral immunity, including isotype during vaccination, particularly by modulating changes in the B cell compartment resulting in a highly non-specific inflammatory B cell response, and a reduction in the vaccine-specific B cell response.

**Fig 5.**
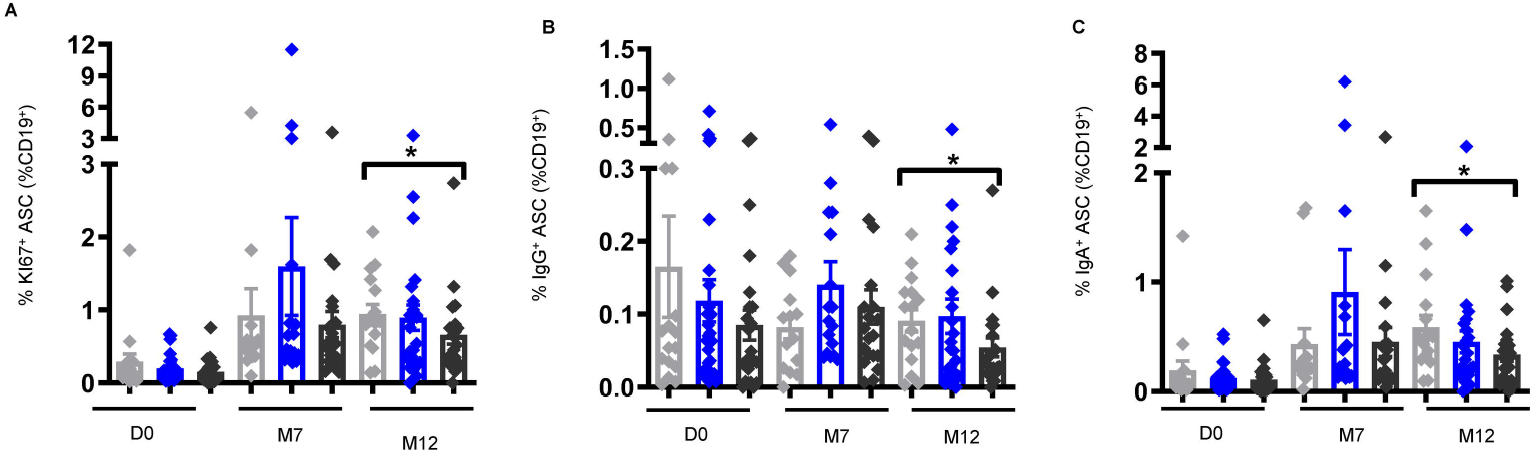
Frequencies of antibody secreting cells (ASCs) are lower at month 12 post-vaccination in individuals with high CAA concentration. Frequencies of (A) KI67^+^ ASCs [CD19^+^CD10^-^IgD^-^ CD71^+^CD38^+^CD20^-^], (B) IgG^+^ ASCs, and (C) IgA^+^ ASCs, were identified by flow cytometry of PBMCs from individuals pre-vaccination (D0) non-infected, n =16, low CAA, n =29, and high CAA, n =24, M7 post-vaccination non-infected, n =14, low CAA, n =17, and high CAA, n =20, and M12 post-vaccination non-infected, n =16, low CAA, n =23, and high CAA, n =21. Data shown as ± SEM. * P ≤ 0.05. Wilcoxon rank-sum test performed on non-infected vs. low CAA, or non-infected vs. high CAA, or low CAA vs. high CAA for each time point separately D0, M7, or M12. Non-infected-light grey, low CAA - blue, and high CAA - dark grey.

### Impact of *S. mansoni* infection on Ig isotypes

B cell isotype switching changes the effector functions of antibodies and improves the effectiveness of the required response to infection or vaccination. Herein, B cell function was further investigated in the context of Ig class-switching by treating PBMCs from non-infected and *S. mansoni*-infected individuals with the TLR9 ligand CpG-ODN 2006 and analyzing the culture supernatant for Ig antibodies using a multiplex bead assay. IgG4, known to be associated with *Schistosoma* reinfection (44), was found to be the only subclass with significant differences between non-infected and *S. mansoni*-infected at pre-vaccination, and had the highest levels in the high worm burden group (no vs. high P=0.002, low vs. high P=0.003) (Fig 6A). We observed similar trends for plasma IgE pre-vaccination (no vs. high P=0.0008, low vs. high P=0.0001); in contrast to CpG-induced IgG4, higher levels were sustained at M12 post-vaccination (no vs. high P=0.001, low vs. high P=0.002) (S6 Fig). Furthermore, a Luminex-based serologic assay was also used to quantify pre-vaccination levels of Schistosoma-specific antibody isotypes and subclasses in the serum of participants, and the mean florescence intensity (MFI) of *S. mansoni*-specific IgG4 was found to be significantly associated with CAA concentration (P=0.0008), with higher levels in individuals with high CAA compared to non-infected (P=0.0003) and individuals with low CAA (P=0.021) (Fig 6B). Post-vaccination, IgA levels in the supernatant of CPG-stimulated PBMCs were found to be significantly different between non-infected and *S. mansoni*-infected individuals, where the lowest levels were observed in individuals with high CAA (no vs. high P=0.031) (Fig 6C). The development of antigen-specific soluble IgA is thought to be critically important in an effective immune response to vaccination (45), and these results reveal that worm burden could influence the host’s ability to mount a specific antibody response.

**Fig 6.**
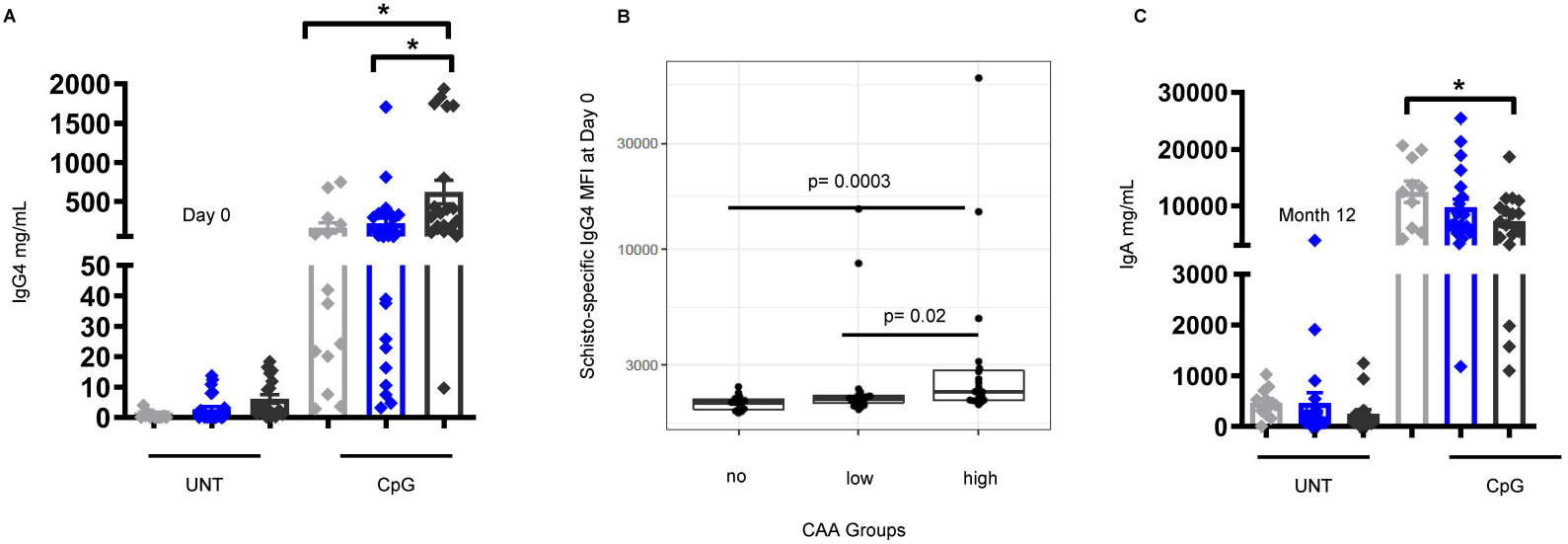
Changes in the Ig isotype profile are evident in *S. mansoni* infection pre- and post-vaccination in response to TLR9 stimulation. (A) IgG4 levels pre-vaccination (D0), and (C) IgA levels at M12 post-vaccination in the culture supernatant of PBMCs stimulated for 7 days with CpG (CpG-ODN [TLR9]) or UNT (untreated). D0 non-infected, n =14, low CAA, n =27, and high CAA, n =21, and M12 post-vaccination non-infected, n =10, low CAA, n =20, and high CAA, n =17. Data shown as ± SEM. * P ≤ 0.05. Wilcoxon matched-pairs signed rank test performed on UNT vs. CpG for each non-infected, or low CAA, or high CAA, and within the CpG-treated group a Wilcoxon rank-sum test on non-infected vs. low CAA, or non-infected vs. high CAA, or low CAA vs. high CAA. Non-infected-light grey, low CAA - blue, and high CAA - dark grey. (B) The mean Florescence intensity (MFI) of serum *S. mansoni*-specific IgG4 at D0 non-infected [no], n =15, low CAA [low], n =21, and high CAA [high], n =20). A student t-test was used to evaluate for the significance of the correlation. P ≤ 0.05 was considered significant.

### Vaccine-Specific Memory CD4^+^ T cells positively correlate with Hepatitis B antibody titers

The CD4^+^ T cell memory compartment houses both the cTfh and Treg populations, which showed variations in their frequencies in instances of *Schistosoma* infection pre- and post-vaccination (Fig 4). To determine the impact of *S. mansoni* infection on HepB-specific memory T cells, PBMCs from non-infected and *S. mansoni*-infected individuals at M7 were treated with a Hepatitis B long envelope protein (HBV LEP) peptide, and CFSE^-^ cells analyzed by flow cytometry were identified as vaccine-specific. Vaccine-specific memory CD4^+^ T and CD8^+^ T cells were significantly higher after peptide stimulation compared to control (DMSO), and lower frequencies of proliferating CD4^+^ T cells, but not CD8^+^ T cells were observed in *S. Mansoni*-infected individuals compared to non-infected (Fig 7A and 7B). Interestingly, and in assessing an association with HepB titers, a positive association was observed between vaccine-specific memory CD4^+^ T cells and HepB titers (t=1.85, P=0.073) (Fig 7C), with a concurrent significant negative correlation between vaccine-specific memory CD8^+^ T cells and HepB titers (t=-2.36, P=0.025) (Fig 7D). These results highlight how CD4^+^ and CD8^+^ memory T cells may play separate and antagonistic roles in their contribution to vaccination responses during infection.

**Fig 7.**
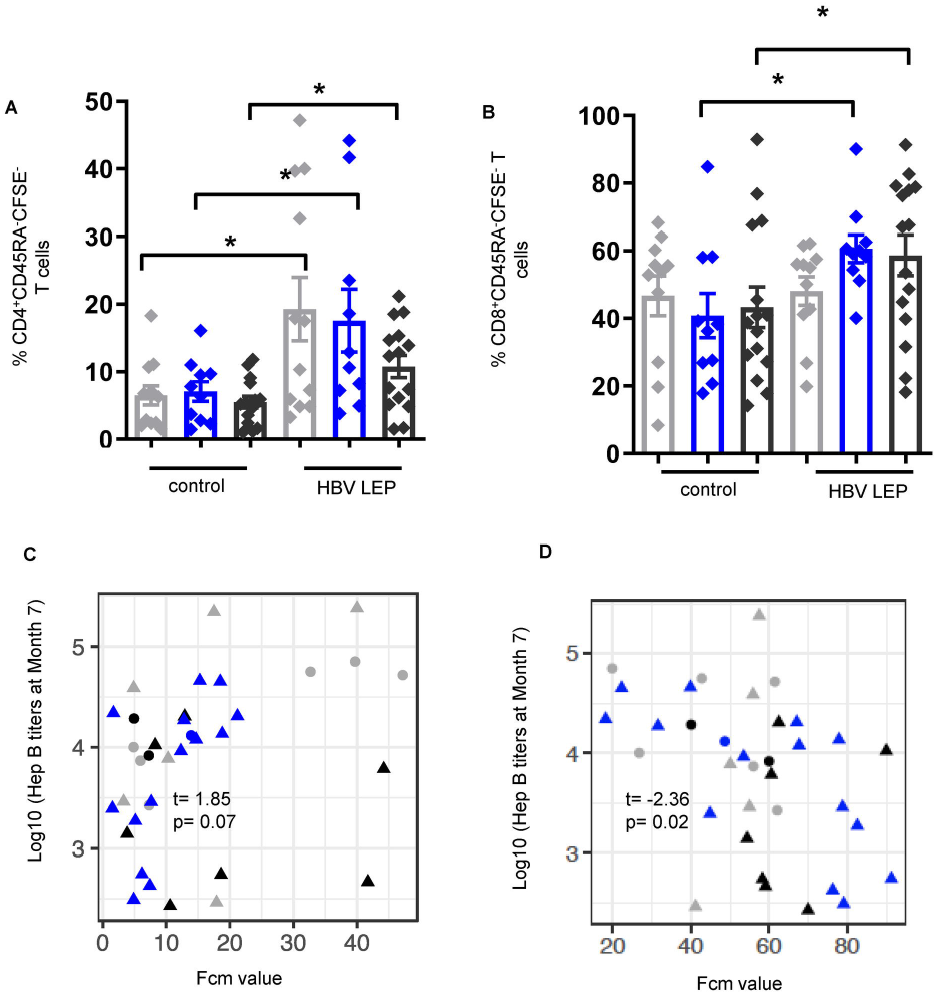
Hepatitis B peptide-specific memory CD4^+^ T cells positively correlate with Hepatitis B titers at month 7 post-vaccination, with a concurrent negative correlation for Hepatitis B peptide-specific memory CD8^+^ T cells. Frequencies of CFSE^-^ (A) CD4^+^CD45RA^-^ memory T (CD4^+^ mem), and (B) CD8^+^CD45RA^-^ memory T cells (CD8^+^ mem), were identified by flow cytometry of PBMCs from individuals at M7 post vaccination non-infected, n =12, low CAA, n =10, and high CAA, n =15, that were stimulated for 6 days with hepatitis B long envelope protein peptide (HBV LEP) or DMSO control (control). Data shown as ± SEM. * P ≤ 0.05. Wilcoxon matched-pairs signed rank test performed on control vs. HBV LEP for each non-infected, or low CAA, or high CAA, and within the HBV-LEP-treated group a Wilcoxon rank-sum test on non-infected vs. low CAA, or non-infected vs. high CAA, or low CAA vs. high CAA within the CpG-treated. Non-infected-light grey, low CAA - blue, and high CAA - dark grey. (C) Linear regressions were fit between Hepatitis B titers and CFSE^-^ (C) CD4^+^ mem, and (D) CD8^+^ mem T cell populations adjusted for sex and student t-tests were used to evaluate for the significance of the association. t (t-statistic). P ≤ 0.05 was considered significant. (Shape: triangle-Male, circle-Female; color: grey-non-infected, black-low CAA, blue-high CAA).

### Monocyte function is positively associated with effective Hepatitis B vaccine responses

Our analysis has pointed to the importance of key cytokines/chemokines involved in innate responses to infection that was detected pre-vaccination, and how they may play a crucial role in modulating the immune response post-vaccination in non-infected and *S. mansoni*-infected individuals with various levels of worm burden. To additionally investigate the importance of innate responses, the frequencies of monocyte populations were determined by flow cytometry of PBMCs from non-infected and *S. mansoni*-infected participants (S7 Fig), however no differences between CAA groups were observed pre-vaccination for all three monocyte populations (S8A-S8C Fig), even though CD14^+^CD16^-^ classical monocytes were found to negatively associate with HepB titers significantly at M7 (coef=-0.027, P=0.017) (S8D Fig). To further elucidate the relationship between monocyte function and HepB vaccine responses, THP-1 cells (a monocyte cell line) were used to carry out an antibody-dependent cellular phagocytosis (ADCP) assay, and ADCP was found to positively correlate with HepB titers at M12 (P=0.0008) (Fig 8A), HepB surface antigen (HbsAg)-specific IgG1 titers (P=0.009) (Fig 8B), and binding of HbsAg-specific antibodies to the cytotoxic Fc gamma receptor 3A (FcγR3A) (P=0.001) (Fig 8C), indicating the importance of monocyte function in an effective humoral immune response to the Hepatitis B vaccine. As we did not observe any associations between monocyte subsets and CAA concentration, suggesting that monocyte frequency is not directly affected by *S. mansoni* infection, we thought it was important to further examine associations of innate immune function with CAA concentration by exploring the induction of cytokine/chemokines.

**Fig 8.**
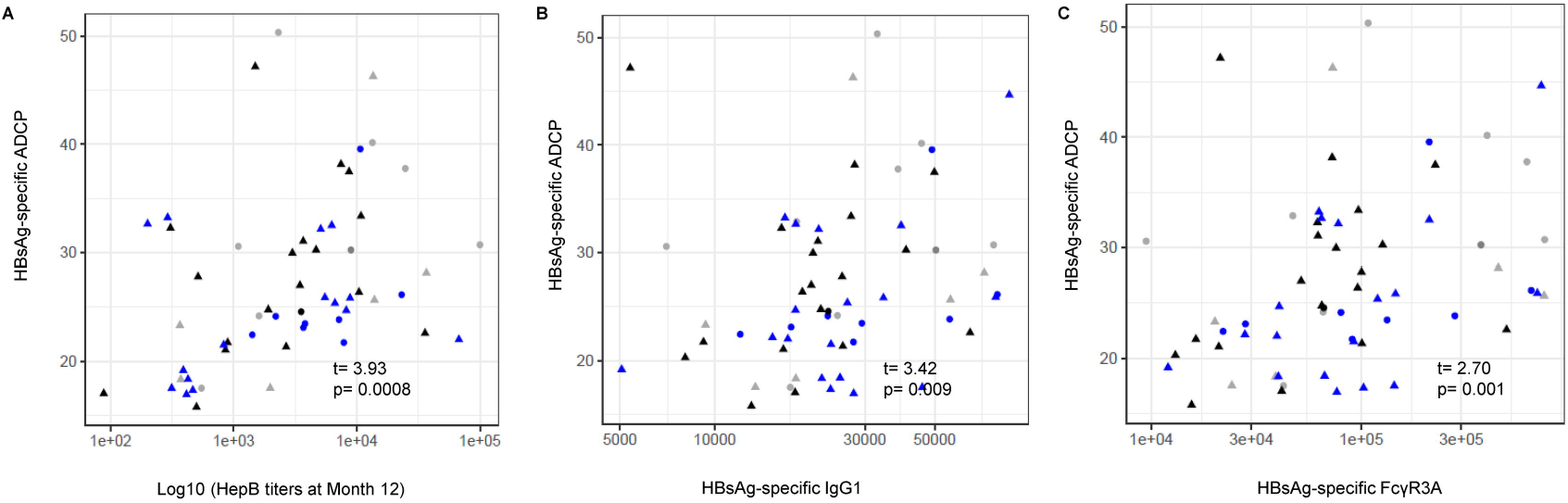
Monocyte function is important in and significantly correlate with Hepatitis B vaccine responses. Antibody-dependent cellular phagocytosis (ADCP) assays with serum from individuals pre-vaccination, non-infected, n =14, low CAA, n =20, and high CAA, n =18, were conducted with cells from the monocyte cell line THP-1. Scatter plots show HBsAg-specific ADCP associations with (A) Hepatitis B titers at M12, and with (B) HBsAg-specific IgG1, and (C) HbsAg-specific antibody binding to FcγR3A (CD16) expression determined by antibody subclass and Fc receptor binding assays using serum samples from pre-vaccinated individuals. Spearman correlation and t-test were used to evaluate for the significance of the correlation. *coef* (regression coefficient). t (t-statistic). P ≤ 0.05 was considered significant. (Shape: triangle-Male, circle-Female; color: grey-non-infected, black-low CAA, blue-high CAA).

### Innate immune-related cytokines/chemokines are significantly lower pre-vaccination and day 12 post-vaccination in instances of high CAA concentration after TLR7/8 stimulation

Toll-like receptors (TLRs) mediate innate immune responses to pathogen-associated molecules (46). To investigate the importance of monocyte function in shaping the response to Hepatitis B vaccination during *S. mansoni* infection, PBMCs from non-infected and *S. mansoni*-infected individuals were stimulated with a viral toll-like receptor (TLR) 7/8 agonist-imidazoquinoline compound CLO97, and the supernatant analyzed by multiplex bead assay for cytokines/chemokines after 18 hours of stimulation. Levels of the CXC chemokine IFN-γ-inducible protein 10 (CXCL10; IP-10), which plays a crucial role in activating innate immune cells, promoting dendritic cell (DC) maturation, and inducing protective T cell responses (47, 48), were significantly lower in CLO97-treated supernatant from the high CAA group pre-vaccination (low vs. high P=0.031) (Fig 9A, S7 Table). Furthermore, supernatant levels of CCL19 (no vs. high P=0.009), CCL26 (no vs. high P=0.014), CCL27 (no vs. high P=0.007), IL-1β (no vs. high P=0.028), and IL-10 (no vs. high P=0.008, low vs. high P=0.053) were also significantly lower in the high CAA group after treatment with CLO97 D12 post-vaccination (Fig 9B-9F, S7 Table). Collectively, these cytokines/chemokines play important roles in early immune responses to viral infection and vaccination including modulation of inflammatory responses and adaptive cell recruitment, activation, and regulation (49–53). These results indicate an association between *S. mansoni* infection and worm burden in the altering of the pre-vaccination, and early post-vaccination innate cytokine environment.

**Fig 9.**
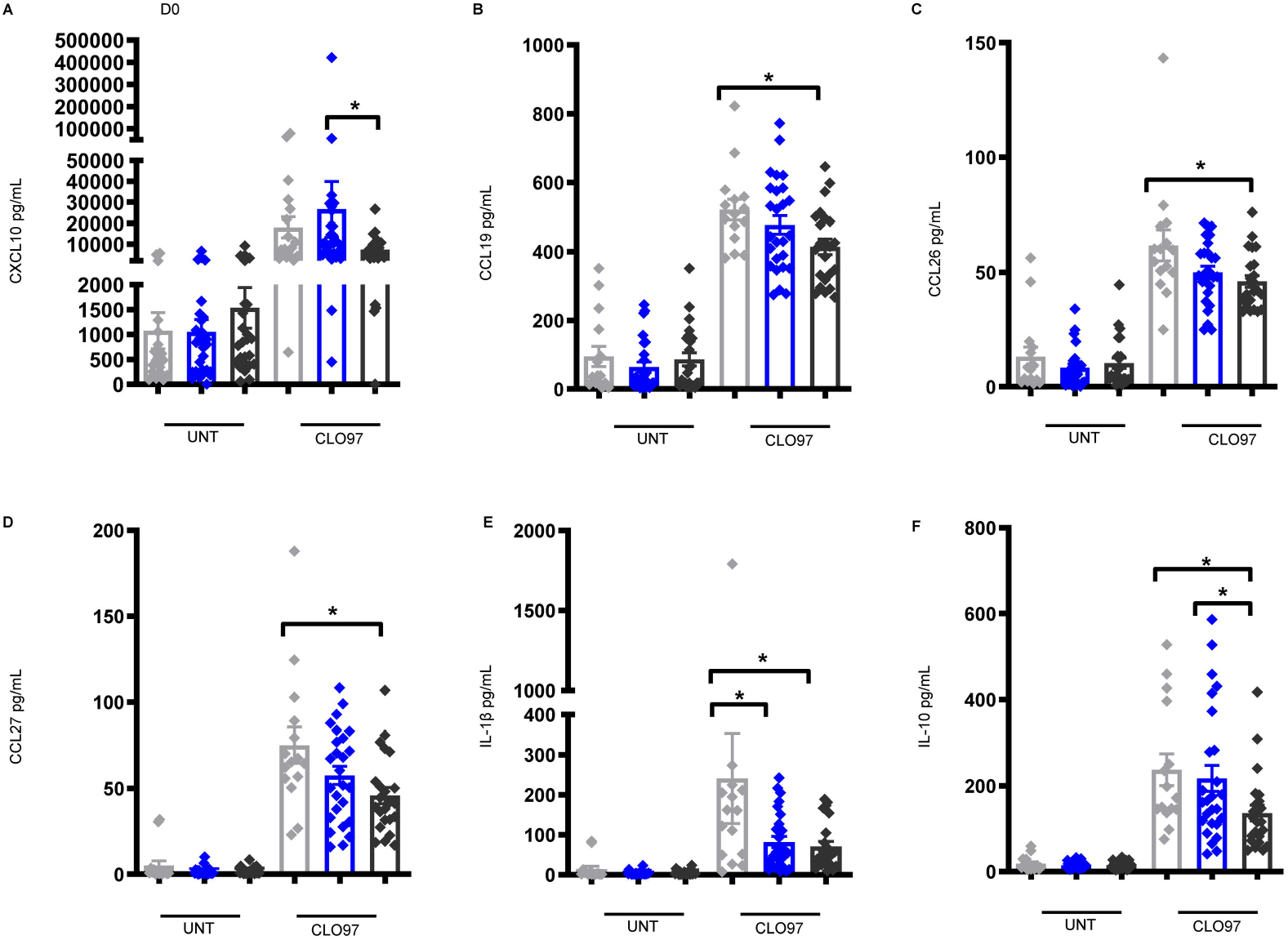
Lower levels of cytokines/chemokines important in the activation and maturation of innate immune cells in *S. mansoni*-infected individuals pre-vaccination and day 12 post-vaccination. (A) CXCL10 levels pre-vaccination (D0) non-infected, n =19, low CAA, n =31, and high CAA, n =25, and (B) CCL19, (C) CCL26, (D) CCL27, (E) IL-1β, and (F) IL-10 levels at D12 post vaccination non-infected, n =15, low CAA, n =26, and high CAA, n =23], in the culture supernatant of PBMCs stimulated for 18 hours with CLO97 (TLR7/8 agonist) or untreated (UNT). Data shown as ± SEM. * P ≤ 0.05 Wilcoxon matched-pairs signed rank test performed on UNT vs. CLO97 or each non-infected, or low CAA, or high CAA, and within the CLO97-treated group a Wilcoxon rank-sum test on non-infected vs. low CAA, or non-infected vs. high CAA, or low CAA vs. high CAA. Non-infected-light grey, low CAA - blue, and high CAA - dark grey.

## Discussion

Vaccination is an essential tool in controlling the spread of infectious diseases, but variations of host immune responses to vaccines across populations (endemic infections, pre-existing infections, geography, biological sex, race/ethnicity) impact the generation of protective immune responses, and thus how vaccines can be designed to treat populations are at greater risk for poor clinical outcomes. These variations in vaccine responses, seen in communities with endemic diseases such as schistosomiasis, have recently been the topic of intense investigation (3, 4). We show that pre-vaccination worm burden specifically influences levels of vaccine-specific antibody production. Individuals from a Ugandan fishing community cohort were stratified into non-infected and *S. mansoni*-infected with low worm burden (low CAA concentration), or high worm burden (high CAA concentration) (33). We show that the pre-vaccination CAA values aligned with groups categorized by low and high worm burden displayed a bimodal distribution, and that increasing CAA concentration was negatively associated with vaccine-specific antibody levels. This association also served as the premise for further investigation to delineate what molecules and cell populations were at play, and responsible for these changes in vaccine-related immune responses post-vaccination.

The major strength of this study lies in this unique cohort, which allowed analysis of individuals pre-vaccination and post-vaccination, with different concentrations of CAA and likely different levels of worm burden from direct exposure, or no *S. mansoni* infection at all; and allowed us to comment definitively on the impact of baseline *S. mansoni* infection on vaccine-induced responses, and more specifically if these induced responses were affected by worm burden. Notably, CAA concentration indicates an active infection and the presence of living worms, allowing for serum CAA concentration to be directly associated with worm burden (27, 28). While egg count data was determined, they are approximately 50% sensitive when done on a single stool sample (54). Furthermore, as the worms themselves do not multiply, egg count data only indicates worm pairs producing eggs and not quantitative data on overall worm burden (55). Our analysis of vaccine-specific antibody titers, cytokines/chemokines, and immune and adaptive cellular responses also allowed us to identify the links between high *S. mansoni* worm burden and lower HepB vaccine responses. While the outcomes of this present study were based solely on the influence of *S. Mansoni* infection on HepB vaccination, we believe the results have the potential to direct other immunization programs in schistosomiasis-impacted communities where repeated exposure is likely.

Our analysis showed induction of several proinflammatory molecules involved in lymphocyte migration and activation, including CCL17 in individuals with high CAA pre-vaccination suggesting a coordinated immune response in instances of high worm burden. Higher levels of CCL17 and sIL-2R showed negative associations with HepB titers in cases of high worm burden, identifying these two molecules as potential biomarkers for inefficient antibody development. CCL17 has been shown to play a key role in recruiting CCR4-expressing T helper and regulatory cells during inflammation (34–36, 38). Furthermore, Wirnsberger and colleagues specifically show that IL-4 induces the expression of CCL17 (56), and we observed higher levels of IL-4 in individuals with high CAA post-vaccination (data not shown) suggesting IL-4 may be playing a role in driving and sustaining the CCL17 responses observed. While T helper 1 and 2 responses, are predominant in Schistosomiasis (14), these chronically skewed environments have been known to be antagonistic to Tfh-B cell interactions that are important for inducing antigen-specific antibody responses (18, 19, 40). Furthermore, alterations in innate immune function in *Schistosoma* infection observed in our cohort, may potentially play a role in these shifts in these cytokine levels; and previous models have also shown that elevation of T-helper cytokines in *S. mansoni* infection correlate specifically with lower vaccine-specific responses to immunization against *Mycobacterium bovis* (BCG) (21), HepB and tetanus toxoid (26).

Several Tfh cell subsets involved in antibody induction during infection and vaccination, including cTfh cells in the periphery, play a key role the generation of high-affinity antibody-producing plasma cells and memory B cells (39). We also now know that changes observed in cTfh profiles are known to reflect the cytokine microenvironment, and specific cTfh subpopulations are known to drive B cell class switching (57). We demonstrated that individuals with high CAA had lower cTfh1 and -2 cells pre-vaccination, which was sustained post-vaccination for cTfh1 and cTfh17 cells in this group. More specifically, a positive association between HepB-specific memory CD4^+^ T cell responses and HepB-specific antibody production was observed. These data highlight the vital role of HepB-specific memory CD4^+^ T helper cells in driving antibody responses, and that changes in this compartment directly influences vaccine responses. In parallel, we also reported higher frequencies of in Foxp3^+^Tregs in individuals with high CAA post-vaccination, suggesting a polarization towards higher frequencies of Tregs. These data strongly suggest that in instances of high worm burden, robust regulatory responses can be induced by key pro-Treg cytokines pre-vaccination, such as CCL17 (36, 37), and other cytokines such as IL-2 which we also observe to be elevated pre-vaccination (S1E Fig), can inhibit Tfh responses (58). We believe this environment at baseline can dictate varied changes to the T cell profile and kinetics post-vaccination and skew the ratio of Tregs: cTfh cells when there is a high worm burden. Interestingly, it has also been shown that the spleen is a major source of Tregs in hepatosplenic schistosomiasis (59), and we can hypothesize that higher frequencies of Tregs in this condition, could contribute to a diminished vaccine-induced response.

In support of our findings, Yin et. al. recently reported that lower responders to HepB vaccination had lower frequencies of cTfh cells and antibody-secreting plasmablasts (42); and Musaigwa and colleagues describe that *S. mansoni* infection specifically induces cell death in bone marrow plasmablasts and plasma cells (60). We show in addition to a lower frequency of cTfh cells in individuals with high CAA, a significant increase in activated B cells but a reduction in proliferating (antigen-specific) ASCs, ASCs expressing IgG and IgA, and that IgA^+^ASCs positively correlate with HepB titers. In addition to these data, we see a significant reduction in TLR9-induced IgA levels post-vaccination. In combination, these results point to a direct role for high worm burden environments pre-vaccination in influencing changes in the immune landscape polarizing it away from cTfh and B cell responses which would hamper vaccine-specific antibody production post-vaccination. Induction of IgA is a significant and desirable aim of vaccination for its advantageous role in mucosal responses and has been proven to be associated with protection effects post-vaccination (45, 61). We have also previously shown that changes to the cytokine microenvironment, particularly hyperinflammation, drives sustained dysregulation of the Tfh-B cell interaction and hence B cell function (40, 41). Together these results emphasize that for successful vaccination, an overall efficient Tfh and B cell response is required.

Innate responses play critical role in shaping the adaptive immune response to vaccines. We show the importance of monocyte function in innate immune responses and desired vaccine responses to HepB. While our analysis showed higher levels of several innate proinflammatory cytokines in the plasma of individuals with high CAA, we did not observe an association between frequencies of monocyte populations and *S. mansoni* infection. However, after stimulation with a viral TLR 7/8 ligand, we show that the levels of induced cytokines varied with *S. mansoni* infection, evidenced by mixed responses pre- and 12 days post-vaccination. These findings give some indication on how pro-inflammatory innate environments could direct the disruption of efficient vaccine-specific responses during *Schistosoma* infection. Furthermore, these results suggest that innate cells are playing a distinctive functional role in changing this early cytokine/chemokine microenvironment, which we know impacts overall adaptive immunity. Specifically, induction of CXCL10 (IP-10), which plays a crucial role in the activation and recruitment of leukocytes and monocytes, promoting DC maturation and inducing protective T cell responses (47, 48), was decreased in individuals with high worm burden pre-vaccination. CXCL10 has been shown to promote the recruitment of CXCR3^+^ cells, which not only include activated T and B cells (62, 63), but also the B-helper Tfh cells important in efficient responses to infection and vaccination (64–66). A CXCL10– inclusive signature was also associated with effective immune responses to a SARS-CoV-2 vaccine after the 1st vaccination (67), and recent vaccination studies also highlighted a link between CXCL10 levels in the serum and innate responses associated with increased vaccine-specific antibody titers (68, 69). Our data, combined with other studies, highlight a vital role for CXCL10 in modulating the early response to vaccination.

Notably, it has been demonstrated that deworming can enhance the host immune response to vaccination (25), and some studies have shown that PZQ treatment can partially restore vaccine-induced immune responses (60, 70). While all enrolled individuals in this current study were treated with PZQ at D12 post-vaccination (Fig 1), it is possible that *Schistosoma*-related effects on baseline immune responses may have already occurred. Furthermore, the rate of reinfection and the extent to which the initial infection was cleared, is also unknown; and as this study also indicates that several proinflammatory cytokines at baseline remain elevated at month 12 post-vaccination, it is possible that individuals with a higher baseline CAA concentration get more reinfections with time, creating a cycle of high CAA concentration driving inefficient immune responses to secondary antigen. Despite these limitations, we observed differences in vaccine-induced responses in individuals with varying pre-vaccinated levels of CAA, irrespective of reported PQZ treatment. A similar observation was seen in an animal study of chronic schistosomiasis and HIV vaccination where schistosomiasis suppressed vaccine responses, and this was maintained regardless of anti-helminth treatment (71). While we know anti-helminth treatment is effective at clearing worms, results suggest that *Schistosoma* infection in this study is associated with irreversible immunomodulatory effects that can influence a host’s response to unrelated antigen, such as those in vaccines.

In this study, we did not evaluate for transgenerational exposure to *Schistosoma*. Previous studies have shown that parasite antigen and maternal antibodies can transfer from mothers to babies in utero and during breastfeeding (72), and some studies have showed lower vaccine responses in the newborns of *Schistosoma*-infected mothers (73–75). These observations highlight the potential for different baseline immune responses in individuals with transgenerational exposure to *Schistosoma* infection and its effect on vaccine responses needs further investigation. It is also likely that some individuals are genetically more susceptible to *Schistosoma* and related hepatic fibrosis than others (76), and further understanding is needed on how susceptibility and high worm load may contribute to how these individuals mount effective vaccine responses. In this study we adjusted for age and sex but people with *Schistosoma* may be more likely to have other exposures (including other worm species, different microbiome) (77), which might influence vaccine response.

In summary, high worm burden in *S. mansoni* infection is associated with the dysregulation of vaccine-induced responses by the cooperation of key cytokines/chemokines that lead to elevated inflammation, and lower levels of cytokines that promote the frequency and function of innate and adaptive immune subsets important in generating vaccine-specific antibodies. Furthermore, a high CCL17 and low CXCL10-inclusive cytokine/chemokine signature pre-vaccination in instances of high worm burden, is also identified. The results from our study could be applied to instruct ongoing and future projects to benefit vaccination programs in *Schistosoma*- and other helminth-endemic communities, particularly in the design of studies that address specific hypotheses such as recently published study protocols from Nkurunungi and colleagues which describe the effect of a more intensive intervention with anti-helminth treatment on vaccine responses in adolescents in Uganda (78, 79).

## Methods

### Study participants and design

Healthy adult volunteers who were enrolled in this study were one-arm of a community-based prospective investigation that took place in fishing communities located in Entebbe, Uganda, called the Stimulated Vaccine Efficacy Trial (SiVET). SiVET was set up to identify populations and assess the implementation of efficacy trial procedures. Hepatitis B and typhoid vaccines were selected as simulated vaccines due to the potential benefit to the fishing communities. Participants aged 18 to 49 years, males and non-pregnant females were enrolled starting in 2015 from one mainland lakeshore fishing community and one island community along Lake Victoria, in Wakiso district. (S1 Table). Inclusion criteria encompassed HIV-uninfected, negative Hep B surface antigen (HbsAg) and core antibody tests, capability, and willingness to provide written informed consent to receive HBV vaccine, and consent for follow-up leading up to 12 months after the first study immunization. Volunteers were not prescreened for active malaria and latent TB infections. Samples from 75 volunteers were available to be used for this present sub-study with the objective to explore predictors of immune responses, (including parasite burden, microbiome, effect of diet on microbiome) and to explore novel methods to identify predictors of vaccine response and pathways activated in effective vaccine immune responses. Adults were screened up to 6 weeks before the first study injection of the Hepatitis B vaccine ENGERIX-B (GlaxoSmithKline Biologicals, derived from recombinant subunit Hepatitis B surface antigen adsorbed on aluminum hydroxide) followed by two booster doses given at months 1 and 6 (Fig 1). Doses were given intramuscularly in the deltoid muscle at 1mL volume containing 20 µg of the vaccine. Whole blood and plasma were collected pre-vaccination (D0), post-1^st^ injection (D3, D7, and D12), and post-booster injections on the day the infections were administered (M1 and M6) (Fig 1). Volunteers were treated at day 12 with Praziquantel (PZQ) as appropriate, for egg counts detected in stool samples at enrollment, D3, and/or D7 (Fig 1). *Schistosoma-*infected participants received 40mg/kg body weight single dose (average 2.4 g) of PZQ.

### Treatment of *S. mansoni*-infected enrolled individuals

Stool samples were collected from enrollees’ pre-vaccination (D0), prior to the first dose of the HepB vaccine, and at D3 and D7 post-vaccination. *Schistosoma* egg counts were subsequently conducted and if stool sample/s were positive, PZQ treatment was administered at D12 post-vaccination. Egg count results were available within one week, after which any infected individuals were treated.

### Circulating anodic antigen (CAA) assay

The circulating anodic antigen assay (CAA) was performed pre-vaccination (Day 0) on serum samples from enrolled participants to reveal ongoing *Schistosoma* infection (29–32). Human negative serum (Sanquin, blood donors, the Netherlands) was spiked with a known concentration of CAA and dilutions made up to eight standard points to provide an appropriate standard series. An extra negative serum sample was included as a duplicate negative control. Samples were evaluated using the SCAA500 protocol with a lower limit of detection threshold of 3pg/mL; 500 mL serum and standards were extracted with an equal volume of 4% w/v trichloroacetic acid (TCA; Merck Life Science NV, the Netherlands), vortexed and incubated at ambient temperature for five minutes. Thereafter, samples and standards were briefly vortexed and spun at 13000g for five minutes and 0.5 mL of clear supernatant was concentrated to 20 mL using Amicon Ultra-0.5 Centrifugal Filter Units devices with a molecular weight cut-off of 10 kDa (Merck Life Science N.V., the Netherlands). Note, for samples with insufficient serum volumes the SCAA20 test format was used which requires only 20uL serum and no concentration step but has a lower limit of detection threshold of 30pg/mL. The resulting TCA soluble fraction (20μL) was added to wells containing 100 ng dry UCP particles (80) (400 nm Y_2_O_2_S:Yb^3+^,Er^3+^) coated with mouse monoclonal anti-CAA antibodies (32) hydrated with 100 mL of high salt lateral flow buffer (HSFS: 200 mM Tris pH8, 270 mM NaCl, 0.5% (v/v) Tween-20, 1% (w/v) BSA. After incubated for one hour at 37°C while shaking at 900rpm the CAA lateral flow strips (32) were placed in the wells, and samples allowed to flow. The strips were then dried overnight and analyzed using an Upcon reader (Labrox Oy, Turku, Finland). The test line signals (T; relative fluorescent units, peak area) were normalized to the flow control signals (FC) of the individual strips and the results expressed as Ratio value (R=T/FC).

### Whole blood processing and sample storage

PBMCs isolation/storage: Whole Blood collected in NaHeparin tube (BD, NJ, USA) was layered over 20ml of Histopaque (Sigma-Aldrich, Darmstadt, Germany) and centrifuged at 400g for 40 minutes at room temperature with no centrifuge brakes. Lymphocytes, platelets, and monocytes found at the plasma-separating medium interface (buffy coat) were recovered and washed in Hanks Balanced Salt solution to remove contaminating platelets, separation media and plasma. The resulting PBMCs (lymphocytes and monocytes) were resuspended in complete RPMI media (RPMI 1640 media supplemented with 10% Fetal Bovine Serum (FBS), 10mM Hepes buffer, 2mM L-glutamine, 1mM sodium pyruvate and 1X penicillin-streptomycin) for counting and concentrated at 10million cells/mL/ vial in FBS with 10% DMSO, frozen down using a rate-controlled freezer and stored in liquid nitrogen.

### Serum separation

Specific vacutainers for serum (SST, Plymouth UK) were invert twice to mix blood and then centrifuged at 1200 g for 10 minutes at room temperature with maximum acceleration and brake after which aliquots into pre-labeled cryovials were made and stored at below -70°C.

### Plasma separation

Plasma from the separation tube of PBMCs was harvested and then centrifuged at 1800 g for 20 minutes at room temperature with maximum acceleration and brake after which aliquots into pre-labeled cryovials without disturbing the platelet pellet at the bottom of the tube and stored at below -70°C.

### Hepatitis B antibody testing

Each serum sample was assayed at screening using three different HBV infection tests for: Hepatitis B surface antigen (HbsAg), Hepatitis B core antibody (anti-HBc), and Hepatitis B surface antibody (anti-HBs). The HbsAg and antiHBc antibodies were tested for using the VIDAS HbsAg Ultra and VIDAS anti-HBc Total II (Biomerieux SA, France) kits respectively on the MinVidas analyzer. The anti-HBs testing was done using the Cobas e 411 analyzer (Roche Diagnostics, Mannheim, Germany). The anti-HBs titer cut off value was 10IU/L. All three tests were used only at screening to differentiate between possible past exposure from active infection, and to ensure only HepB-negative individuals were recruited and administered the vaccine. Measurement of anti-HBs antibody was conducted for all subsequent follow-up visits.

### PBMC culture

PBMCs from non-infected and *S. mansoni*-infected individuals pre-vaccination and post-vaccination were thawed and rested for 3 hours before being placed into culture. Samples were used for ex-vivo immunostaining and flow cytometry analysis, and for TLR stimulation assays. PBMCs were suspended in RPMI medium supplemented with L-glutamine (Corning Cellgro, Manassas, VA, USA), 10% FBS and 1 X [50 U] penicillin-streptomycin (Invitrogen, Carlsbad, CA, USA) (R10F). 1 x10^6^ cells were added to 5-ml polypropylene tubes for CLO97 stimulation (Imidazoquinoline Compound; TLR7/8 agonist) (Invivogen, San Diego, CA, USA), and 0.5 x 10^6^ cells added to the wells of 96-well U-bottom plates for CpG-ODN 2006 stimulation (TLR9 agonist) (Invivogen). Optimal concentration for CLO97 stimulation was selected as previously described (81). CpG-ODN 2006 was titrated on CFSE-labeled PBMCs, and optimal TLR9 activity assessed on proliferating CD19^+^ cells by flow cytometry. Cells were allowed to rest at 37°C under a 5% CO^2^ atmosphere for 3 hours prior to addition of TLR agonists at the following concentrations: CLO97 (0.5ug/ml) and CpG (1ug/ml). Cells were cultured also at 37°C under a 5% CO^2^ atmosphere for 18hrs (CLO97), and 7 days (CpG-ODN 2006). After culture, PBMCs were collected and washed in preparation for analysis by multiplex immunoassays or flow cytometry. For HBV peptide stimulation, PBMCs were first labeled with CFSE using a Cell Trace™ CFSE cell proliferation kit (ThermoFisher Scientific, Waltham, MA, USA), and rested for 3 hours in the conditions described above. In a 96-well deep well plate (USA Scientific, Ocala, FL, USA), 2 x 10^6^ cells in 1mL RPMI medium supplemented with 8% human serum (Access Biologicals, Vista, CA, USA), 1% penicillin-streptomycin and 10ng/mL IL-2 (Miltenyi Biotec, Auburn, CA, USA) (R8H-IL-2) were stimulated with either Pepmix™ HBV (Large envelope protein) (JPT Peptide Technologies, Berlin, Germany) at 1ug/mL or control (R8H-IL-2 with 0.2% DMSO) on Day 1 and cultured in the conditions described above. On Day 3, cells were supplemented with fresh R8H-IL-2 by half media replacement; and on Day 6, PBMCs were collected and washed in preparation for measurement of immune recall responses by flow cytometry.

### Cytokine and chemokine analysis (multiplex immunoassay)

Plasma collected from whole blood and supernatant collected from stimulated PBMCs were analyzed for chemokines/cytokines and Ig isotypes using magnetic bead multiplex assays. The following human analyte premixed panels were used: Bio-Plex human chemokine panel (Bio-Rad, Hercules, CA, USA): I-309 (CCL1), MCP-1 (CCL2), MIP-1α (CCL3), MCP-3 (CCL7), MCP-2 (CCL8), Eotaxin (CCL11), MCP-4 (CCL13), MIP-1α (CCL15), TARC (CCL17), MIP-3β (CCL19), 6Ckine (CCL21), MIP-3α (CCL20), MDC (CCL22), MPIF-1 (CCL23), Eotaxin-2 (CCL24), TECK (CCL25), Eotaxin-3 (CCL26), CTACK (CCL27), GM-CSF, GRO-α (CXCL1), GRO-β (CXCL2), ENA-78 (CXCL5), GCP-2 (CXCL6), MIG (CXCL9), IP-10 (CXCL10), I-TAC (CXCL11), SDF-1A+β (CXCL12), BCA-1 (CXCL13), SCYB16 (CXCL16), Fractalkine (CX3CL1), MIF, IL-1β, IL-2, IL-4, IL-6, IL-8, IL-10, IL-16, TNF-α, and IFN-γ. Custom ProcartaPlex 34-plex (ThermoFisher Scientific): APRIL, BAFF, CD30, CD40L, G-CSF, IFN-A, IL-12P70, IL-13, IL-15, IL-16, IL-17A, IL-18, IL-1A, IL-20, IL-21, IL-22, IL-23, IL-27, IL-2R, IL-3, IL-31, IL-5, IL-7, IL-9, LIF, M-CSF, TNF-R2, TNF-B, TRAIL, TSLP, TWEAK, VEGF-A, IFNB, and IL-29/IFN-lambda1. Bio-Plex human isotype panel (Bio-Rad): IgM, IgA, IgG1, IgG2, IgG3, IgG4. IgE was measured using Bio-Plex human IgE isotype assay (Bio-Rad). The manufacturer’s protocol was followed. Data was acquired on a Bio-Plex 200 system (using bead regions defined in the protocol) and analyzed with the Bio-Plex Manager 6.1 software (Bio-Rad).

### Flow cytometry analysis of PBMCs

For ex-vivo analysis, 1 x 10^6^ PBMCs per well were incubated with Fixable aqua viability stain 405 (ThermoFisher Scientific) to discriminate dead from live cells then stained with antibody panels described (S8 and S9 Tables). To detect intracellular expression of markers, cells were fixed and permeabilized using the Foxp3/Transcription factor staining buffer set (ThermoFisher Scientific) and immunostained using FOXP3-FITC (clone, PCH101) (ThermoFisher Scientific) (S8 Table) and KI67-FITC (BD Biosciences, San Jose, CA, USA) (S9 Table). Following HBV peptide stimulation, PBMCs were stained with the surface antibody panel described (S10 Table), then stained with 7-AAD (BD Biosciences) for 10 minutes prior to analysis to discriminate dead from live cells. Flow cytometry measurements were made on a BD LSRFortessa™ (BD Biosciences) and collected data analyzed using FlowJo software version 10.7.1 (BD).

### Antibody-dependent cellular phagocytosis (ADCP) assay

An antibody-dependent cellular phagocytosis (ADCP) assay to investigate monocyte function was conducted. HbsAg was biotinylated using an EZ-Link™ Sulfo-NHS-LC-Biotinylation Kit (ThermoFisher Scientific) and coupled to FluoSpheres™ NeutrAvidin™-Labeled Microspheres (ThermoFisher Scientific). To form immune complexes, antigen-coupled beads were incubated for 2 hours at 37°C with 1:25 diluted serum samples and then washed to remove unbound immunoglobulin. After washing, immune complexes were incubated for 16-18 hours with a monocyte cell line, THP-1 cells [25,000 THP-1 cells per well at a concentration of 1.25×105 cells/ml in RPMI (ThermoFisher Scientific) + 10% FBS (Sigma-Aldrich), and following incubation, cells were fixed with 4% paraformaldehyde (Poly Scientific R&D, Bay Shore, NY, USA). Percentage of fluorosphere positive cells was analyzed on a LSRII flow cytometer (BD Biosciences). The phagocytosis score was defined.

### Antibody subclass and Fc receptor binding

HbSAg-specific antibody subclass titers and Fc receptor binding profiles were analyzed with a custom multiplex Luminex assay as described previously (82). In brief, HbSAg was coupled to magnetic Luminex beads (Luminex Corp, TX, USA). Coupled beads were incubated with diluted serum samples, washed, and IgG subclasses detected with a 1:100 diluted PE-conjugated secondary antibody for IgG1 (clone: HP6001; Southern Biotech, AL, USA). For the FcγR binding, a respective PE–streptavidin (Agilent Technologies, Santa Clara, CA, USA) coupled recombinant and biotinylated human FcγR protein was used as a secondary probe.

### Statistics

Statistical inference evaluating the differences of HepB titers between non-infected versus low CAA, non-infected versus high CAA and low CAA versus high CAA was determined using Wilcoxon rank-sum test. Wilcoxon rank-sum tests were performed on non-infected vs. low CAA, or non-infected vs. high CAA, or low CAA vs. high CAA within each time point for plasma Luminex assays, and ex-vivo flow cytometry analysis. Pairwise Wilcoxon signed rank test were performed on untreated vs. treated for each non-infected, or low CAA, or high CAA for CpG, HBV-LEP and CLO97 stimulation assays; and within the treated groups, a Wilcoxon rank-sum test was conducted on non-infected vs. low CAA, or non-infected vs. high CAA, or low CAA vs. high CAA. Linear regression analyses were adjusted for sex (no significant association between age and HepB titers) and student t-tests were used to evaluate for the significance of the association. Sensitivity analyses were performed using the R package sensemakr (83) used to assess the robustness of the results of the linear regression analysis to an unobserved confounding variable. Benjamini-Hochberg adjustment was used to control for multiple testing for each assay. P ≤ 0.05 was considered significant for all analyses.

### Study approval

Informed consent was obtained from all participants prior to being enrolled in the studies. All studies were approved by the Uganda Virus Research Institute Research Ethics Committee, reference number GC/127/15/07/439 and the Uganda National Council of Science and Technology, reference number HS 1850.

## Author contributions

RM, TM, and SF share first authorship. RM and TM are first and second co-authors respectively, as they conceptualized, designed, and performed experiments, analyzed data, and wrote the manuscript. SF is the third co-author and analyzed data and wrote the manuscript. AK, NK, A Nanvubya, PK, BSB and YM designed the SiVET clinical study. AK, AE, PK, JKL and BSB conceptualized the SiVET study. YB, JKL, GC, GA, EM, BO, TN, IN, A Namuniina, AS, JM, PKK, VMB, BSB, NK, JN, WS, CJDD, PB and MO performed experiments and analyzed data. PLAMC and GJVD designed clinical assays and PLAMC edited the manuscript. EM, BO, TN, IN, AN, AS, JM, PKK, and NK managed enrollee recruitment and participation in the study. PF, MAP, AME, and RPS provided conceptual advice and edited the manuscript. EKH conceptualized experiments and wrote the manuscript. All authors reviewed and approved the manuscript. RM and EKH had final responsibility for the decision to submit for publication.

## Supporting information captions

**S1 Fig. Elevated plasma cytokines/chemokines in *S. mansoni*-infected individuals pre-vaccination.** (A) CCL7, (B) CCL11, (C) CCL22, (D) CCL24, (E) IL-2, and (F) M-CSF levels in the plasma of non-infected, n = 19, low CAA, n = 32, and high CAA, n = 24, individuals pre-vaccination (day 0). Data shown as ± SEM. * P ≤ 0.05. Wilcoxon rank-sum test performed on non-infected vs low CAA, or non-infected vs high CAA, or low CAA vs high CAA for each time point separately. Non-infected-light grey, low CAA - blue, and high CAA - dark grey.

**S2 Fig. CCL17 and soluble IL-2R levels in individuals with *S. mansoni* infection correlate with Hepatitis B titers post-vaccination.** Scatter plots of Hepatitis B titers at M12 as a function of (A-B) CCL17 and (D-E) soluble IL-2R cytokine levels in the plasma of non-infected participants at D0, n =19, low CAA, n =32, and high CAA, n =24, and M12 post-vaccination, n =15, low CAA, n =29, and high CAA, n =19. P ≤ 0.05. Linear regressions were fit between Hepatitis B titers and the cytokines adjusted for sex and student t-tests were used to evaluate for the significance of the association. t (t-statistic). P ≤ 0.05 was considered significant. (Shape: circle-females, triangle-males; color: grey-non-infected, black-low CAA, blue-high CAA). (C) Soluble IL-2R levels in the plasma of non-infected individuals pre-vaccination (D0), n =19, low CAA, n =32, and high CAA, n =24, and M12 post-vaccination, n =15, low CAA, n =29, and high CAA, n =19. Non-infected-light grey, low CAA - blue, and high CAA - dark grey.

**S3 Fig. Flow cytometry gating strategy for T cell populations.** Representative plots showing non-circulating T follicular helper cells (cTfh) populations as CD3^+^CD4^+^CD45RA^-^CD25^-^CXCR5^-^ and cTfh populations as CD3^+^CD4^+^CD45RA^-^CD25^-^CXCR5^+^, identifying cTfh1 as CXCR3^+^, cTfh2 as CXCR3^-^ CCR6^-^, and cTfh17 as CXCR3^-^CCR6^+^. Regulatory T cells (Tregs) were identified as CD3^+^CD4^+^CD45RA^-^CD127^-^CD25^+^Foxp3^+^.

**S4 Fig. Flow cytometry gating strategy for B cell populations.** Representative plots showing activated B cells (ABC) as CD19^+^CD10^-^IgD^-^CD71^+^CD38^-^CD20^+^ and antibody secreting B cells (ASC) as CD19^+^CD10^-^IgD^-^CD71^+^CD38^+^CD20^-^.

**S5 Fig. Frequencies of Activated B cells (ABC) were elevated in *S. mansoni* infection pre- and post-vaccination.** Frequencies of (A) ABCs [CD19^+^CD10^-^IgD^-^CD71^+^CD38^-^CD20^+^], and (B) IgG^+^ ABCs, were identified by flow cytometry of PBMCs from individuals pre-vaccination (D0) non-infected, n = 16, low CAA, n = 29, and high CAA, n = 24], M7 post-vaccination non-infected, n = 14, low CAA, n = 17, and high CAA, n = 20, and M12 post-vaccination non-infected, n = 16, low CAA, n = 23, and high CAA, n = 21. Data shown as ± SEM. * P ≤ 0.05. Wilcoxon rank-sum test performed on non-infected vs low CAA, or non-infected vs high CAA, or low CAA vs high CAA for each time point separately D0, M7, or M12. Non-infected-light grey, low CAA - blue, and high CAA - dark grey. (C) Linear regressions were fit between Hepatitis B titers and IgA^+^ ASCs adjusted for sex and student t-tests were used to evaluate for the significance of the association. t (t-statistic). P ≤ 0.05 was considered significant. (Shape: triangle-Male, circle-Female; color: grey-non-infected, black-low CAA, blue-high CAA).

**S6 Fig. Plasma IgE is elevated pre-vaccination and remains elevated at month 12 post-vaccination in *S. mansoni*-infected individuals.** Plasma IgE levels pre-vaccination (D0) and M12 post-vaccination. D0 non-infected, n = 17, low CAA, n = 20, and high CAA, n = 19], and M12 post-vaccination non-infected, n = 15, low CAA, n = 26, and high CAA, n = 20. Data shown as ± SEM. * P ≤ 0.05. Wilcoxon rank-sum test performed on non-infected vs low CAA, or non-infected vs high CAA, or low CAA vs high CAA for each time point separately D0, or M12. Non-infected-light grey, low CAA - blue, and high CAA - dark grey.

**S7 Fig. Flow cytometry gating strategy for monocyte populations.** Representative plots showing non-classical (NC) monocytes as CD3^-^CD19^-^CD14^dim^CD16^+^, classical monocytes (CL) as CD3^-^CD19^-^ CD14^+^CD16^+^, and intermediate monocytes (INT) as CD3^-^CD19^-^CD14^+^CD16^-^.

**S8 Fig. Flow cytometry gating strategy for monocyte populations.** Frequencies of monocyte populations (A) (classical [CD3^-^CD19-CD14^+^CD16^-^], (B) intermediate [CD3^-^CD19^-^CD14^+^CD16^+^], and (C) non-classical [CD3^-^CD19^-^CD14^dim^CD16^+^] were identified by flow cytometry of PBMCs from individuals pre-vaccination, non-infected, n =14, low CAA, n =20, and high CAA, n =18. Non-infected-light grey, low CAA - blue, and high CAA - dark grey. (D-F) Scatter plot showing the frequencies of monocytes subsets as a function of Hepatitis B titers at M7 post-vaccination. Linear regressions were fit between Hepatitis B titers and the monocyte subset frequencies adjusted for sex and experiment batch, and student t-tests were used to evaluate for the significance of the association. Spearman correlation and t-test were used to evaluate for the significance of the correlation. *coef* (regression coefficient). t (t-statistic). P ≤ 0.05 was considered significant.

**S1 Table.** Study participant information.

**S2 Table.** Statistical information for plasma Luminex cytokine/chemokine levels.

**S3 Table.** Sensitivity analysis for plasma Luminex cytokine/chemokine levels correlated with Hepatitis B titers.

**S4 Table.** Statistical information for Frequency of T cells.

**S5 Table.** Statistical information for Frequency of B cells.

**S6 Table.** Sensitivity analysis for B cell flow cytometry frequencies correlated with Hepatitis B titers.

**S7 Table.** Statistical information for supernatant cytokine/chemokine levels from CLO97-stimulated PBMCs.

**S8 Table.** Ex vivo T cell staining panel for PBMCs.

**S9 Table.** Ex vivo B cell staining panel for PBMCs.

**S10 Table.** T cell staining panel for PBMCs after Hepatitis B peptide stimulation.

## Data Availability

All data produced in the present study are available upon reasonable request to the authors

## Acknowledgements

This work was supported by NIH funding as part of Human Immune Project Consortium (HIPC) to EKH and RPS #U19 AI128910. The SiVET was supported by a grant from the United States Agency for International Development (USAID) [USAID reference number AID-OAA-A-16-00032]. The contents are the responsibility of the authors and does not necessarily reflect the views of USAID or the United States Government. We are grateful to the study participants, and healthcare and research staff from the Immunomodulation and Vaccines Programme and the Pathogen Genomics Phenotype and Immunity Programme at the MRC/UVRI and LSHTM Uganda Research Unit, the UVRI-IAVI HIV Vaccine Program at the College of Health Sciences at Makerere University, Kampala-Uganda, and the International AIDS Vaccine initiative, for the invaluable contribution to this study.

