## Supplementary material for "*Schistosoma mansoni* infection alters the host pre-vaccination environment resulting in blunted Hepatitis B vaccination immune responses": S1 Table

**S1 Table. Study participant information**

| **Variable** | **Total** | **Non-infected** | **Low CAA** | **High CAA** | **P value** |
| --- | --- | --- | --- | --- | --- |
| **Volunteers enrolled with D0 HepB Ab titers and CAA, n** | 79^AB^ | 19 | 35 | 25 |  |
| **Average age (range)** | 28 (18-46) | 30 (23-46) | 29 (18-46) | 27 (18-45) | 0.278 |
| **Sex, male/female (female %)** | 55/24 (30%) | 9/10  (52.6%) | 22/13  (37.1%) | 24/1  (4.0%) | 0.005 |
| **D0 CAA value, pg/mL (average)** | 217 | 1 | 23 | 698 |  |
| **M7 HepB Ab titers, n participants** | 64 | 16 | 26 | 22 |  |
| **M7 HepB Ab titers, pg/mL (range)** | 31847  (2-240000) | 48930  (291-240000) | 35089  (2-184300) | 15589  (310-75900) | 0.347 |
| **M12 HepB Ab titers, n participants** | 64 | 16 | 27 | 21 |  |
| **M12 HepB Ab titers, pg/mL (range)** | 8234  (2-100000) | 13881  (366-100000) | 7468  (2-67510) | 4916  (88.1-36830) | 0.585 |

^A^4 donor samples were unavailable for analysis in this study. ^B^Participants included in the study were HBsAg, anti-HBc and anti-HBs antibody negative as part of the inclusion criteria. D0- pre-vaccination, M7- month 7 post-vaccination, M12, month 12 post-vaccination. P ≤ 0.05 was considered significant. Wilcoxon rank-sum test.
