## Supplementary material for "*Schistosoma mansoni* infection alters the host pre-vaccination environment resulting in blunted Hepatitis B vaccination immune responses": S2 Table

**S2 Table. Statistical information for plasma Luminex cytokine/chemokine levels**.

| Analyte | Timepoint | P no vs low | P no vs high | P low vs high |
| --- | --- | --- | --- | --- |
| TARC (CCL17) | Day 0 | 0.0018 | <0.0001 | 0.113 |
| MIP-3β (CCL19) | Day 0 | 0.0035 | <0.0001 | 0.0374 |
| CXCL9 | Day 0 | 0.0118 | <0.0001 | 0.0022 |
| IL-2R | Day 0 | 0.8916 | 0.0777 | 0.0736 |
| CCL7 | Day 0 | 0.0073 | 0.0031 | 0.7633 |
| CCL11 | Day 0 | 0.1329 | 0.0206 | 0.0728 |
| CCL22 | Day 0 | 0.0354 | 0.0047 | 0.0998 |
| CCL24 | Day 0 | 0.0423 | 0.0328 | 0.9572 |
| IL-2 | Day 0 | 0.013 | 0.0022 | 0.1346 |
| M-CSF | Day 0 | 0.131 | 0.3512 | 0.0079 |
| TARC (CCL17) | Month 12 | 0.5748 | 0.0164 | 0.3746 |
| MIP-3β (CCL19) | Month 12 | 0.5834 | 0.0426 | 0.1339 |
| CXCL9 | Month 12 | 0.1532 | 0.0035 | 0.0309 |
| IL-2R | Month 12 | 0.2006 | 0.1167 | 0.9202 |

Non-infected [no], low CAA [low], and high CAA [high]. Wilcoxon rank-sum test performed on non-infected vs low CAA, or non-infected vs high CAA, or low CAA vs high CAA within each time point Day 0 or Month 12. P ≤ 0.05.
