## Supplementary material for "*Schistosoma mansoni* infection alters the host pre-vaccination environment resulting in blunted Hepatitis B vaccination immune responses": S3 Table

**S3 Table. Sensitivity analysis for plasma Luminex cytokine/chemokine levels correlated with Hepatitis B titers**

|  | Outcome: log10 (HepB titers at Month 12) | | | | | |
| --- | --- | --- | --- | --- | --- | --- |
| Cytokine | **Est.** | **S.E.** | **t-value** | $\mathbf{R}_{\boldsymbol{Y\sim D\vert X}}^{\mathbf{2}}$ | $\mathbf{RV}_{\mathbf{q=1}}$ | $\mathbf{RV}_{\boldsymbol{q=1,\alpha=0.05}}$ |
| CCL17 at Day 0 | -0.0175 | 0.00580 | -3.01 | 13.7% | 32.7% | 12.3% |
| CCL17 at Month 12 | -0.00645 | 0.00269 | -2.40 | 9.79% | 28.0% | 4.99% |
| IL-2R at Day 0 | -0.000130 | 5.85e-5 | -2.22 | 10.7% | 29.2% | 2.67% |
| IL-2R at Month 12 | -7.97e-5 | 2.70e-5 | -2.95 | 13.9% | 32.9% | 11.8% |

Sensitivity analysis for the multivariate regression model with HepB titers at Month 12 as dependent variables and cytokine expression and sex as independent variables. $R_{Y\sim D|X}^{2}$ is the total amount of variance of HepB titers an unobserved confounder needs to explain for the cytokine effect to be zero. $\mathrm{RV}_{q=1}$ is the robustness value (the amount of residual variance an unobserved confounder needs to explain) for bringing the cytokine estimate to zero. $\mathrm{RV}_{q=1,\alpha=0.05}$ is the robustness value (the amount of residual variance an unobserved confounder needs to explain) for bringing the lower bound of the 95% confidence interval to zero. Est.: estimator; S.E.: standard error.
