## Supplementary material for "*Schistosoma mansoni* infection alters the host pre-vaccination environment resulting in blunted Hepatitis B vaccination immune responses": S4 Table

**S4 Table. Statistical information for Frequency of T cells**.

| T cell population | Timepoint | P no vs low | P no vs high | P low vs high |
| --- | --- | --- | --- | --- |
| cTfh1 | Day 0 | 0.2193 | 0.3378 | 0.0228 |
| cTfh2 | Day 0 | 0.8315 | 0.0138 | 0.014 |
| cTfh17 | Day 0 | 0.8007 | 0.055 | 0.0904 |
| CXCR5^-^ Treg | Day 0 | 0.8625 | 0.6179 | 0.444 |
| non cTfh | Day 0 | 0.5824 | 0.3319 | 0.5808 |
| cTfh1 | Month 7 | 0.6883 | 0.1558 | 0.0311 |
| cTfh2 | Month 7 | 0.4928 | 0.5686 | 0.0694 |
| cTfh17 | Month 7 | 0.7467 | 0.4209 | 0.1191 |
| CXCR5^-^ Treg | Month 7 | 0.2108 | 0.0385 | 0.4021 |
| non cTfh | Month 7 | 0.2108 | 0.4945 | 0.4194 |
| cTfh1 | Month 12 | 0.6475 | 0.0297 | 0.0035 |
| cTfh2 | Month 12 | 0.5075 | 0.6258 | 0.1715 |
| cTfh17 | Month 12 | 0.0236 | 0.0058 | 0.3225 |
| CXCR5^-^ Treg | Month 12 | 0.5709 | 0.0465 | 0.1605 |
| non cTfh | Month 12 | 0.8749 | > 0.9999 | 0.8749 |

Non-infected [no], low CAA [low], and high CAA [high]. Wilcoxon rank-sum test performed on non-infected vs low CAA, or non-infected vs high CAA, or low CAA vs high CAA within each time point Day 0, Month 7, or Month 12. P ≤ 0.05. Circulating T follicular helper cell (cTfh); Regulatory T cell (Treg).
