## Supplementary material for "*Schistosoma mansoni* infection alters the host pre-vaccination environment resulting in blunted Hepatitis B vaccination immune responses": S5 Table

**S5 Table. Statistical information for Frequency of B cells**.

| B cell population | Timepoint | P no vs low | P no vs high | P low vs high |
| --- | --- | --- | --- | --- |
| ABC | Day 0 | 0.0936 | 0.0483 | 0.5377 |
| IgG^+^ AB | Day 0 | 0.1256 | 0.0154 | 0.2842 |
| KI67^+^ ASC | Day 0 | 0.6095 | 0.128 | 0.2351 |
| IgG^+^ ASC | Day 0 | >0.9999 | 0.4157 | 0.3966 |
| IgA^+^ ASC | Day 0 | 0.9766 | 0.2406 | 0.2211 |
| ABC | Month 7 | 0.0233 | 0.0386 | 0.7922 |
| IgG^+^ ABC | Month 7 | 0.0406 | 0.0321 | 0.8389 |
| KI67^+^ ASC | Month 7 | 0.4865 | 0.9244 | 0.5118 |
| IgG^+^ ASC | Month 7 | 0.183 | 0.6976 | 0.4104 |
| IgA^+^ ASC | Month 7 | 0.7616 | 0.9931 | 0.5614 |
| ABC | Month 12 | 0.2305 | 0.0396 | 0.518 |
| IgG^+^ ABC | Month 12 | 0.2536 | 0.0325 | 0.33 |
| KI67^+^ ASC | Month 12 | 0.4581 | 0.0396 | 0.4046 |
| IgG^+^ ASC | Month 12 | 0.5816 | 0.0461 | 0.366 |
| IgA^+^ ASC | Month 12 | 0.1487 | 0.0288 | 0.5882 |

Non-infected [no], low CAA [low], and high CAA [high]. Wilcoxon rank-sum test performed on non-infected vs low CAA, or non-infected vs high CAA, or low CAA vs high CAA within each time point Day 0, Month 7, or Month 12. P ≤ 0.05. Activated B cell (ABC); Antibody secreting cell (ASC).
