## Supplementary material for "*Schistosoma mansoni* infection alters the host pre-vaccination environment resulting in blunted Hepatitis B vaccination immune responses": S6 Table

**S6 Table. Sensitivity analysis for B cell flow cytometry frequencies correlated with Hepatitis B titers**

|  | Outcome: log10 (HepB titers at Month 12) | | | | | |
| --- | --- | --- | --- | --- | --- | --- |
| Subset | **Est.** | **S.E.** | **t-value** | $\mathbf{R}_{\boldsymbol{Y\sim D\vert X}}^{\mathbf{2}}$ | $\mathbf{RV}_{\mathbf{q=1}}$ | $\mathbf{RV}_{\boldsymbol{q=1,\alpha=0.05}}$ |
| % IgA^+^ ASC at Month 7 | -0.0764 | 0.0940 | -0.813 | 1.54% | 11.8% | 0% |
| % IgA^+^ ASC at Month 12 | 0.479 | 0.228 | 2.10 | 7.82% | 25.2% | 1.01% |

Sensitivity analysis for the multivariate regression model with HepB titers at Month 12 as dependent variables and the percentage of IgA^+^ ASC in CD19^+^ cells and sex as independent variables. $R_{Y\sim D|X}^{2}$ is the total amount of variance of HepB titers an unobserved confounder needs to explain for the immune subset effect to be zero. $\mathrm{RV}_{q=1}$ is the robustness value (the amount of residual variance an unobserved confounder needs to explain) for bringing the immune subset estimate to zero. $\mathrm{RV}_{q=1,\alpha=0.05}$ is the robustness value (the amount of residual variance an unobserved confounder needs to explain) for bringing the lower bound of the 95% confidence interval to zero. Est.: estimator; S.E.: standard error.
