## Supplementary material for "*Schistosoma mansoni* infection alters the host pre-vaccination environment resulting in blunted Hepatitis B vaccination immune responses": S7 Table

**S7 Table. Statistical information for supernatant cytokine/chemokine levels from CLO97-stimulated PBMCs**.

| Analyte | Timepoint | P no vs low | P no vs high | P low vs high |
| --- | --- | --- | --- | --- |
| CXCL10 | Day 0 | 0.7665 | 0.1711 | 0.0314 |
| CCL19 | Day 12 | 0.3582 | 0.0092 | 0.084 |
| CCL26 | Day 12 | 0.092 | 0.0139 | 0.1963 |
| CCL27 | Day 12 | 0.256 | 0.0074 | 0.1326 |
| IL-1β | Day 12 | 0.0522 | 0.028 | 0.8817 |
| IL-10 | Day 12 | 0.4488 | 0.0082 | 0.0526 |

Non-infected [no], low CAA [low], and high CAA [high]. Wilcoxon rank-sum test performed on CLO97-treated non-infected vs low CAA, or non-infected vs high CAA, or low CAA vs high CAA within each time point Day 0 or Day 12. P ≤ 0.05.
