## Supplementary material for "*Schistosoma mansoni* infection alters the host pre-vaccination environment resulting in blunted Hepatitis B vaccination immune responses": S8 Table

**S8 Table. Ex vivo T cell staining panel for PBMCs**

| Antibody | Color | Clone | Company |
| --- | --- | --- | --- |
| CD3 | AF700 | HIT3a | Biolegend |
| CD4 | APCCy7 | RPA-T4 | Biolegend |
| CD45RA | ECD | 2H4LDH11LDB9 | Beckman Coulter |
| CXCR5 | AF647 | RF8 B2 | BD Biosciences |
| CXCR3 | BV421 | G025H7 | Biolegend |
| CCR6 | PE | 11A9 | BD Biosciences |
| CD8 | BV650 | RPA-T8 | Biolegend |
| PD-1 | PECy7 | EH2.2H7 | Biolegend |
| CD25 | PECY5 | B696 | Biolegend |
| CD38 | BV605 | HIT2 | Biolegend |
| CD127 | PercpCy5.5 | A01905 | Biolegend |
| LIVE/DEAD | Amcyan |  | Thermofisher |
| FOXP3 | FITC | PCH101 | Thermofisher |
