## Supplementary material for "*Schistosoma mansoni* infection alters the host pre-vaccination environment resulting in blunted Hepatitis B vaccination immune responses": S9 Table

**S9 Table. Ex vivo B cell staining panel for PBMCs**

| Antibody | Color | Clone | Company |
| --- | --- | --- | --- |
| CD19 | AF700 | HIB19 | Biolegend |
| CD20 | BV650 | 2H7 | Biolegend |
| CD21 | V450 | B-ly4 | BD Biosciences |
| CD27 | PE | O323 | Biolegend |
| IgD | AF647 |  | Southern Biotech |
| IgM | PECY5 | G20-127 | BD Biosciences |
| IgG | PECy7 | G18-145 | BD Biosciences |
| IgA | AF594 |  | Jackson Immno |
| CD10 | APCCy7 | HI10A | Biolegend |
| CD71 | Percp/Cy5.5 | CY1G4 | Biolegend |
| CD38 | BV605 | HIT2 | Biolegend |
| LIVE/DEAD | Amcyan |  | Thermofisher |
| KI67 | FITC | B56 | BD Biosciences |
