## Supplementary material for "*Schistosoma mansoni* infection alters the host pre-vaccination environment resulting in blunted Hepatitis B vaccination immune responses": S10 Table

**S10 Table. T cell staining panel for PBMCs after Hepatitis B peptide stimulation**

| Antibody | Color | Clone | Company |
| --- | --- | --- | --- |
| CD3 | A700 | HIT3a | Biolegend |
| CD4 | APCCY7 | RPA-T4 | Biolegend |
| CD8 | PECY7 | RPA-T8 | Biolegend |
| CD45RA | ECD | 2H4LDH11LDB9 | BD Biosciences |
| CFSE | FITC |  | Thermofisher |
| 7AAD | PECY5 |  | BD Biosciences |
